# Impact of COVID-19 and Effects of Vaccination with BNT162b2 on Patient-Reported Health-Related Quality of Life, Symptoms, and Work Productivity Among US Adult Outpatients with SARS-CoV-2

**DOI:** 10.1101/2022.08.31.22279264

**Authors:** Manuela Di Fusco, Xiaowu Sun, Mary M. Moran, Henriette Coetzer, Joann M. Zamparo, Laura Puzniak, Mary B. Alvarez, Ying P. Tabak, Joseph C. Cappelleri

## Abstract

**Background:** Although there is extensive literature on the clinical benefits of COVID-19 vaccination, data on humanistic effects are limited. This study evaluated the impact of SARS-CoV-2 infection on symptoms, Health Related Quality of Life (HRQoL) and Work Productivity and Impairment (WPAI) prior to and one month following infection, and compared results between individuals vaccinated with BNT162b2 and those unvaccinated.

**Methods:** Subjects with ≥1 self-reported symptom and positive RT-PCR for SARS-CoV-2 at CVS Health US test sites were recruited between 01/31/2022-04/30/2022. Socio-demographics, clinical characteristics and vaccination status were evaluated. Self-reported symptoms, HRQoL, and WPAI outcomes were assessed using questionnaires and validated instruments (EQ-5D-5L, WPAI-GH) across acute COVID time points from pre-COVID to Week 4, and between vaccination groups. Mixed models for repeated measures were conducted for multivariable analyses, adjusting for several covariates. Effect size (ES) of Cohen’s d was calculated to quantify the magnitude of outcome changes within and between vaccination groups.

**Results:** The study population included 430 subjects: 197 unvaccinated and 233 vaccinated with BNT162b2. Mean (SD) age was 42.4 years (14.3), 76.0% were female, 38.8% reported prior infection and 24.2% at least one comorbidity. Statistically significant differences in outcomes were observed compared with baseline and between groups. The EQ-Visual analogue scale scores and Utility Index dropped in both cohorts at Day 3 and increased by Week 4, but did not return to pre-COVID levels. The mean changes were statistically lower in the BNT162b2 cohort at Day 3 and Week 4. The BNT162b2 cohort reported lower prevalence and fewer symptoms at index date and Week 4. At Week 1, COVID-19 had a large impact on all WPAI-GH domains: the work productivity time loss among unvaccinated and vaccinated was 65.0% and 53.8%, and the mean activity impairment was 50.2% and 43.9%, respectively. With the exception of absenteeism at Week 4, the BNT162b2 cohort was associated with statistically significant less worsening in all WPAI-GH scores at both Week 1 and 4.

**Conclusions:** COVID-19 negatively impacted HRQoL and work productivity among mildly symptomatic outpatients. Compared with unvaccinated, those vaccinated with BNT162b2 were less impacted by COVID-19 infection and recovered faster.

## BACKGROUND

The impact of the COVID-19 pandemic on the sustainability of quality of life of patients has been reported globally [1–4]. The prolonged multisystem symptoms associated to SARS-CoV-2 infection can negatively affect daily activities, ability to work, and social interactions, leading to poor health-related quality of life (HRQoL) [1–4].

The introduction of COVID-19 vaccination has significantly impacted the COVID-19 response, and evidence regarding the efficacy, safety and effectiveness of vaccination is extensive [5]. However, there is limited research on the potential benefits of vaccination on physical, mental, social, emotional functioning and economic well-being. Most of the studies assessing humanistic outcomes of COVID-19 infection have been limited to inpatients [1, 2, 6] were conducted outside of the US or focused on specific disease states and organ-specific functions [7–9]. Leveraging a US national retail pharmacy SARS-CoV-2 test database and using validated patient-reported outcome measures (PROMs), this study assessed COVID-19 symptoms, HRQoL and WPAI prior to through one month following SARS-CoV-2 infection in outpatients, and compared results between unvaccinated individuals and those vaccinated with BNT162b2.

## METHODS

### Study Design and Participants

The source population consisted of individuals testing for SARS-CoV-2 at one of ∼5,000 CVS Health test sites across the US. As part of the registration process for scheduling a SARS-CoV-2 test at CVS Health, individuals are required to complete a screening questionnaire including demographics, symptoms, comorbidities, and vaccination status. The screening variables and RT-PCR test results are loaded in an analytic dataset, where ∼80-90% of test results are reported within 2-3 days. Leveraging this analytic platform, this study was designed as a prospective survey-based patient-reported outcomes study targeting adults ≥18 with a positive RT-PCR test result and self-reporting at least one symptom. These individuals were emailed an invitation as soon as the test results became available, no later than 4 days from testing. The email invitation directed the potential participants to an e-consent website to learn about the study, survey schedule and informed consent. Figure 1 summarizes the study design. Recruitment of participants was carried out between 01/31/2022 and 04/30/2022 (Ct.gov NCT05160636).

**Figure 1.**
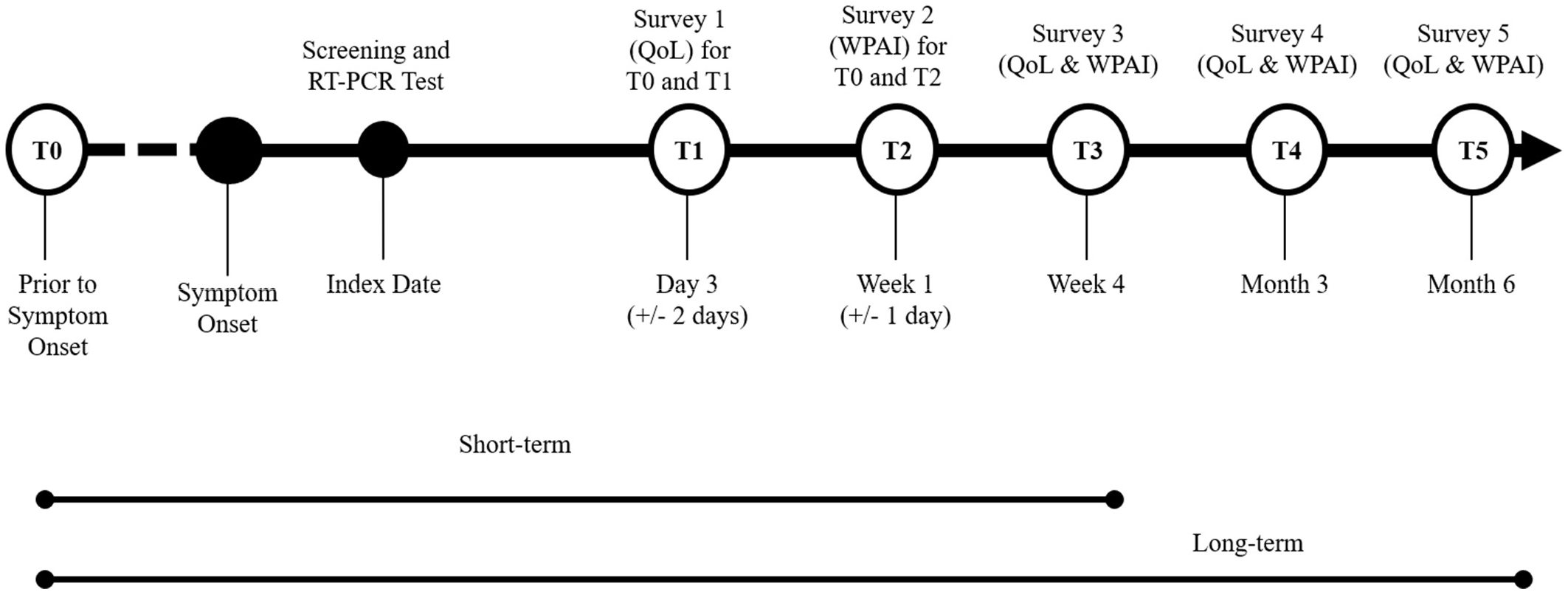
Study design ^a^ ^a^ QoL refers to the EQ-5D-5L survey

### Data Sources and Variables

#### Baseline characteristics and symptoms

Baseline characteristics of the participants were obtained via the CVS Health pre-test screening questionnaire. These included self-reported demographics, comorbidities (including immunocompromised status), COVID-19 vaccination history, social determinants of health including the Social Vulnerability Index (SVI), work and/or residency in a high-risk or healthcare setting, and symptoms. The list of baseline COVID-19 symptoms was based on the CDC [10].

#### Exposure groups

Immunocompetent participants were considered fully vaccinated with BNT162b2 if they self-reported receipt of 2 doses of BNT162b2 ≥ 14 days of SARS-CoV-2 testing. They were considered partially vaccinated if reporting receipt of a single dose and boosted if reporting receipt of 3 doses. Participants self-reporting an immunocompromising condition and receipt of 3 doses were considered fully vaccinated (i.e., 3-dose primary series completion); if reporting 4 doses, they were considered boosted. Participants were considered unvaccinated if they did not report any COVID-19 vaccine dose prior to testing. Heterologous schedules were excluded.

#### HRQoL

To assess HRQoL, we used the validated EQ-5D-5L questionnaire [11, 12]. On the day of enrollment, consented participants completed the EQ-5D-5L questionnaire twice, using two versions: a modified version where all the questions were past tense to retrospectively assess pre-COVID-19 baseline QoL, and the standard version in present tense to assess current QoL. To minimize responder bias, the order of administration of the two versions was random.

Subsequent completion was requested at one month (short-term study design in Figure 1). The EQ-5D-5L results at each time point were converted into the Utility Index (UI) using the US-based weights by Pickard et al [12, 13].

#### Work Productivity and Activity Impairment

To measure impairments in both paid work and unpaid work, we used the Work Productivity and Activity Impairment General Health V2.0 (WPAI:GH) measure [14, 15]. Participants were asked to complete this questionnaire twice, seven days after their RT-PCR test: once referencing seven days prior to COVID-19 symptom onset and an additional assessment referencing the past seven days. Similar to the EQ-5D-5L, subsequent completion of the WPAI was requested at one month (Figure 1). Four WPAI scores were computed at each time point: percent of worktime missed (absenteeism), percent of impairment while working (presenteeism), percent of work productivity loss (considering both absenteeism and presenteeism), and percent of activity impairment. Only employed subjects were included for work productivity analyses.

#### Post-COVID 19 Symptoms and Vaccination Status Update

To supplement the pre-test screening questionnaire and enable the collection of on-going or new symptoms after the acute phase, participants were sent an additional survey four weeks following the test asking to complete a checklist of COVID-19 related symptoms based on the CDC list [16], To confirm vaccination status, participants’ subsequent responses to vaccination date questions were compared with their index responses; if responses did not match, the information was queried and adjudicated, and the latest information was typically used.

### Statistical Analysis

Descriptive statistics were used to analyze participant characteristics at baseline. Continuous variables were described using means and standard deviations. Categorical variables were reported using number and percentage distributions. For continuous variables, t-tests were used to test difference in means and Wilcoxon tests were used to test difference in medians. For categorical variables, chi-square tests were used to test differences between groups When cell frequency was less than 5, Fisher’s exact tests were used for 2-by-2 tables and Freeman-Halton tests for r-by-c tables [17, 18]. P values were all two-sided and not adjusted for multiplicity.

Mixed models for repeated measures (MMRM) [19] were used to estimate the magnitude of COVID-19 impact on HRQoL and WPAI over time. Models included variables of time, self-reported SARS-CoV-2 vaccination status, and interaction of time by vaccination status, as well as covariates of participant pre-COVID-19 symptom onset score, sociodemographic characteristics (age, sex, regions, social vulnerability, race/ethnicity, high risk occupations), previously tested positive for COVID-19, severity of acute illness (number of symptoms reported on index date), and immunocompromised status. Assessment time was fitted as a categorical covariate and a repeated effect (repeated by subject). Least squares mean (LS mean) and standard errors of PRO scores for each time point of assessment were calculated. Per guidelines, no adjustment was made for missing data when scoring the EQ-5D-5L UI and WPAI [11, 15]. Missing data at each time were not imputed. All available data were included in the analysis.

Cohen’s d, or a variation of it, was calculated to assess the magnitude of score change from baseline within the BNT162b2 vaccinated cohort and, separately, the unvaccinated cohort, as well as the difference between BNT162b2 and unvaccinated cohorts [20, 21]. Specifically, within-cohort effect size (ES) was calculated as mean change from baseline to follow-up, divided by the standard deviation of change scores from baseline to follow-up. Between-cohort ES was calculated as the difference in mean changes from baseline between cohorts, divided by the pooled standard deviation of change scores. When calculating model based ESs, the numerators were the predicted mean change from the model for within-cohort ESs, and predicted differences from the model for between-cohort ESs. Denominators were the corresponding observed standard deviations. Values of 0.2, 0.5, and 0.8 standard deviation (SD) units represent small, medium, and large ES, respectively. These cut-off estimates have been widely used to establish important differences in HRQoL studies [22]. As such, we considered the magnitude of (standardized) effect sizes of at least 0.20 SD units as important or meaningful differences in gauging the magnitude of within-patient change and between-group differences. All data obtained were collected and analyzed with SAS Version 9.4 (SAS Institute, Cary, NC). The study followed the Strengthening the Reporting of Observational Studies in Epidemiology (STROBE) reporting guideline [23].

## RESULTS

### Baseline characteristics

A total of 39,889 eligible candidates were outreached. Of those, 676 consented and completed the first survey, for a consent rate of 1.7%. Compared with individuals in the CVS Health analytic dataset who did not participate in our study, the study sample was over-represented by women and Caucasians, with slightly more individuals vaccinated and with comorbidities (Supplemental Table 1). The final study population included 430 subjects (Figure 2). 100% completed the EQ-5D-5L questionnaire at pre-COVID-19 baseline and at Day 3, and 77.0% completed it at Week 4. The WPAI-GH questionnaire was completed by 88.1% of the participants at pre-COVID-19 baseline, 88.1% at Week 1 and 76.9% at Week 4.

**Table 1.**
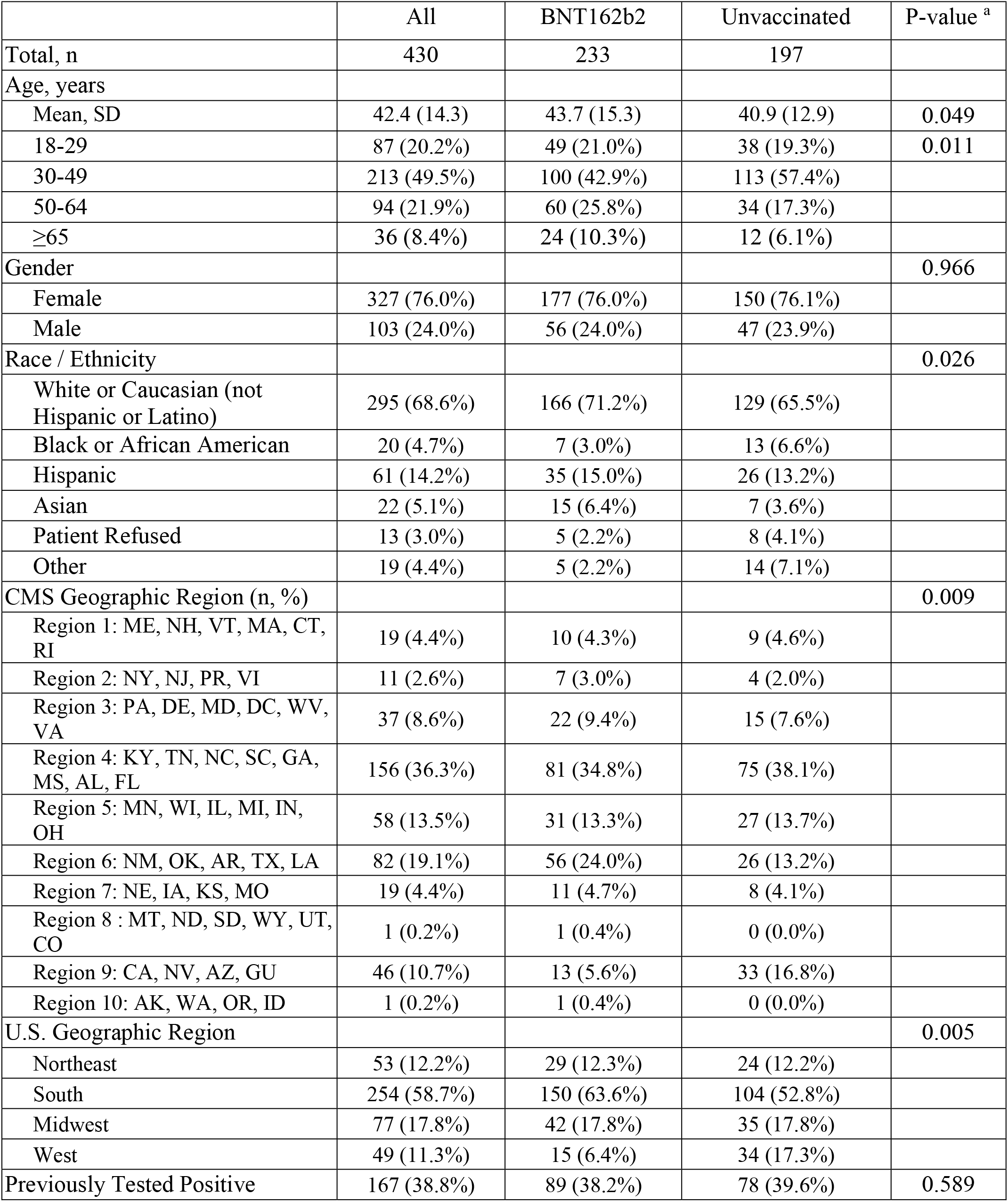
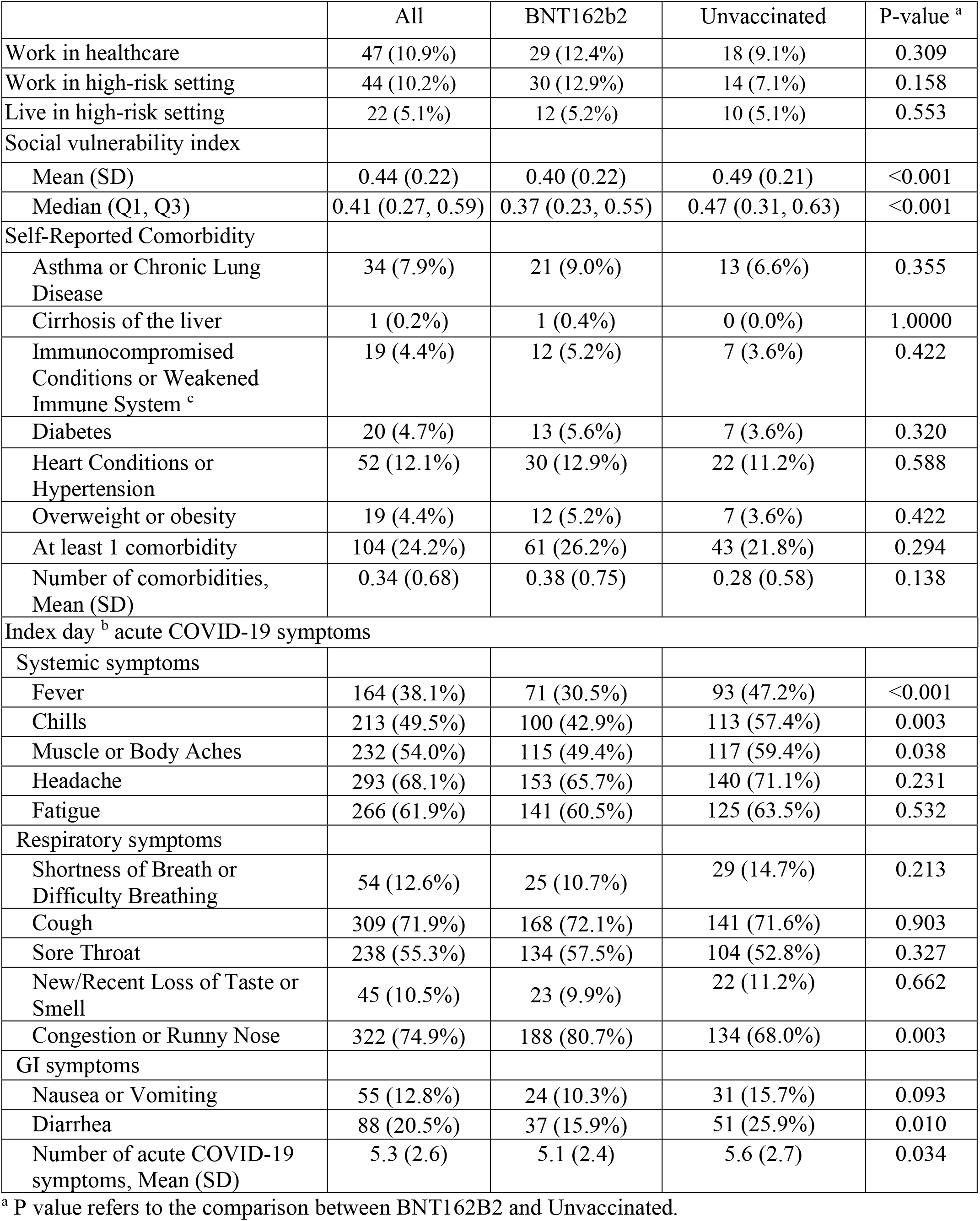
Patient Characteristics on Index Day

|  | All | BNT162b2 | Unvaccinated | P-value <sup>a</sup> |
| --- | --- | --- | --- | --- |
| Total, n | 430 | 233 | 197 |  |
| Age, years |  |  |  |  |
| Mean, SD | 42.4 (14.3) | 43.7 (15.3) | 40.9 (12.9) | 0.049 |
| 18-29 | 87 (20.2%) | 49 (21.0%) | 38 (19.3%) | 0.011 |
| 30-49 | 213 (49.5%) | 100 (42.9%) | 113 (57.4%) |  |
| 50-64 | 94 (21.9%) | 60 (25.8%) | 34 (17.3%) |  |
| ≥65 | 36 (8.4%) | 24 (10.3%) | 12 (6.1%) |  |
| Gender |  |  |  | 0.966 |
| Female | 327 (76.0%) | 177 (76.0%) | 150 (76.1%) |  |
| Male | 103 (24.0%) | 56 (24.0%) | 47 (23.9%) |  |
| Race / Ethnicity |  |  |  | 0.026 |
| White or Caucasian (not Hispanic or Latino) | 295 (68.6%) | 166 (71.2%) | 129 (65.5%) |  |
| Black or African American | 20 (4.7%) | 7 (3.0%) | 13 (6.6%) |  |
| Hispanic | 61 (14.2%) | 35 (15.0%) | 26 (13.2%) |  |
| Asian | 22 (5.1%) | 15 (6.4%) | 7 (3.6%) |  |
| Patient Refused | 13 (3.0%) | 5 (2.2%) | 8 (4.1%) |  |
| Other | 19 (4.4%) | 5 (2.2%) | 14 (7.1%) |  |
| CMS Geographic Region (n, %) |  |  |  | 0.009 |
| Region 1: ME, NH, VT, MA, CT, RI | 19 (4.4%) | 10 (4.3%) | 9 (4.6%) |  |
| Region 2: NY, NJ, PR, VI | 11 (2.6%) | 7 (3.0%) | 4 (2.0%) |  |
| Region 3: PA, DE, MD, DC, WV, VA | 37 (8.6%) | 22 (9.4%) | 15 (7.6%) |  |
| Region 4: KY, TN, NC, SC, GA, MS, AL, FL | 156 (36.3%) | 81 (34.8%) | 75 (38.1%) |  |
| Region 5: MN, WI, IL, MI, IN, OH | 58 (13.5%) | 31 (13.3%) | 27 (13.7%) |  |
| Region 6: NM, OK, AR, TX, LA | 82 (19.1%) | 56 (24.0%) | 26 (13.2%) |  |
| Region 7: NE, IA, KS, MO | 19 (4.4%) | 11 (4.7%) | 8 (4.1%) |  |
| Region 8 : MT, ND, SD, WY, UT, CO | 1 (0.2%) | 1 (0.4%) | 0 (0.0%) |  |
| Region 9: CA, NV, AZ, GU | 46 (10.7%) | 13 (5.6%) | 33 (16.8%) |  |
| Region 10: AK, WA, OR, ID | 1 (0.2%) | 1 (0.4%) | 0 (0.0%) |  |
| U.S. Geographic Region |  |  |  | 0.005 |
| Northeast | 53 (12.2%) | 29 (12.3%) | 24 (12.2%) |  |
| South | 254 (58.7%) | 150 (63.6%) | 104 (52.8%) |  |
| Midwest | 77 (17.8%) | 42 (17.8%) | 35 (17.8%) |  |
| West | 49 (11.3%) | 15 (6.4%) | 34 (17.3%) |  |
| Previously Tested Positive | 167 (38.8%) | 89 (38.2%) | 78 (39.6%) | 0.589 |
| Work in healthcare | 47 (10.9%) | 29 (12.4%) | 18 (9.1%) | 0.309 |
| Work in high-risk setting | 44 (10.2%) | 30 (12.9%) | 14 (7.1%) | 0.158 |
| Live in high-risk setting | 22 (5.1%) | 12 (5.2%) | 10 (5.1%) | 0.553 |
| Social vulnerability index |  |  |  |  |
| Mean (SD) | 0.44 (0.22) | 0.40 (0.22) | 0.49 (0.21) | <0.001 |
| Median (Q1, Q3) | 0.41 (0.27, 0.59) | 0.37 (0.23, 0.55) | 0.47 (0.31, 0.63) | <0.001 |
| Self-Reported Comorbidity |  |  |  |  |
| Asthma or Chronic Lung Disease | 34 (7.9%) | 21 (9.0%) | 13 (6.6%) | 0.355 |
| Cirrhosis of the liver | 1 (0.2%) | 1 (0.4%) | 0 (0.0%) | 1.0000 |
| Immunocompromised Conditions or Weakened Immune System <sup>c</sup> | 19 (4.4%) | 12 (5.2%) | 7 (3.6%) | 0.422 |
| Diabetes | 20 (4.7%) | 13 (5.6%) | 7 (3.6%) | 0.320 |
| Heart Conditions or Hypertension | 52 (12.1%) | 30 (12.9%) | 22 (11.2%) | 0.588 |
| Overweight or obesity | 19 (4.4%) | 12 (5.2%) | 7 (3.6%) | 0.422 |
| At least 1 comorbidity | 104 (24.2%) | 61 (26.2%) | 43 (21.8%) | 0.294 |
| Number of comorbidities, Mean (SD) | 0.34 (0.68) | 0.38 (0.75) | 0.28 (0.58) | 0.138 |
| Index day <sup>b</sup> acute COVID-19 symptoms |  |  |  |  |
| Systemic symptoms |  |  |  |  |
| Fever | 164 (38.1%) | 71 (30.5%) | 93 (47.2%) | <0.001 |
| Chills | 213 (49.5%) | 100 (42.9%) | 113 (57.4%) | 0.003 |
| Muscle or Body Aches | 232 (54.0%) | 115 (49.4%) | 117 (59.4%) | 0.038 |
| Headache | 293 (68.1%) | 153 (65.7%) | 140 (71.1%) | 0.231 |
| Fatigue | 266 (61.9%) | 141 (60.5%) | 125 (63.5%) | 0.532 |
| Respiratory symptoms |  |  |  |  |
| Shortness of Breath or Difficulty Breathing | 54 (12.6%) | 25 (10.7%) | 29 (14.7%) | 0.213 |
| Cough | 309 (71.9%) | 168 (72.1%) | 141 (71.6%) | 0.903 |
| Sore Throat | 238 (55.3%) | 134 (57.5%) | 104 (52.8%) | 0.327 |
| New/Recent Loss of Taste or Smell | 45 (10.5%) | 23 (9.9%) | 22 (11.2%) | 0.662 |
| Congestion or Runny Nose | 322 (74.9%) | 188 (80.7%) | 134 (68.0%) | 0.003 |
| GI symptoms |  |  |  |  |
| Nausea or Vomiting | 55 (12.8%) | 24 (10.3%) | 31 (15.7%) | 0.093 |
| Diarrhea | 88 (20.5%) | 37 (15.9%) | 51 (25.9%) | 0.010 |
| Number of acute COVID-19 symptoms, Mean (SD) | 5.3 (2.6) | 5.1 (2.4) | 5.6 (2.7) | 0.034 |

<sup>a</sup> P value refers to the comparison between BNT162B2 and Unvaccinated.
<sup>b</sup> COVID-19 test nasal swab day
<sup>c</sup> Immunocompromised conditions includes compromised immune system (such as from immuno-compromising drugs, solid organ or blood stem cell transplant, HIV, or other conditions), conditions that result in a weakened immune system, including cancer treatment, and kidney failure or end stage renal disease

**Figure 2.**
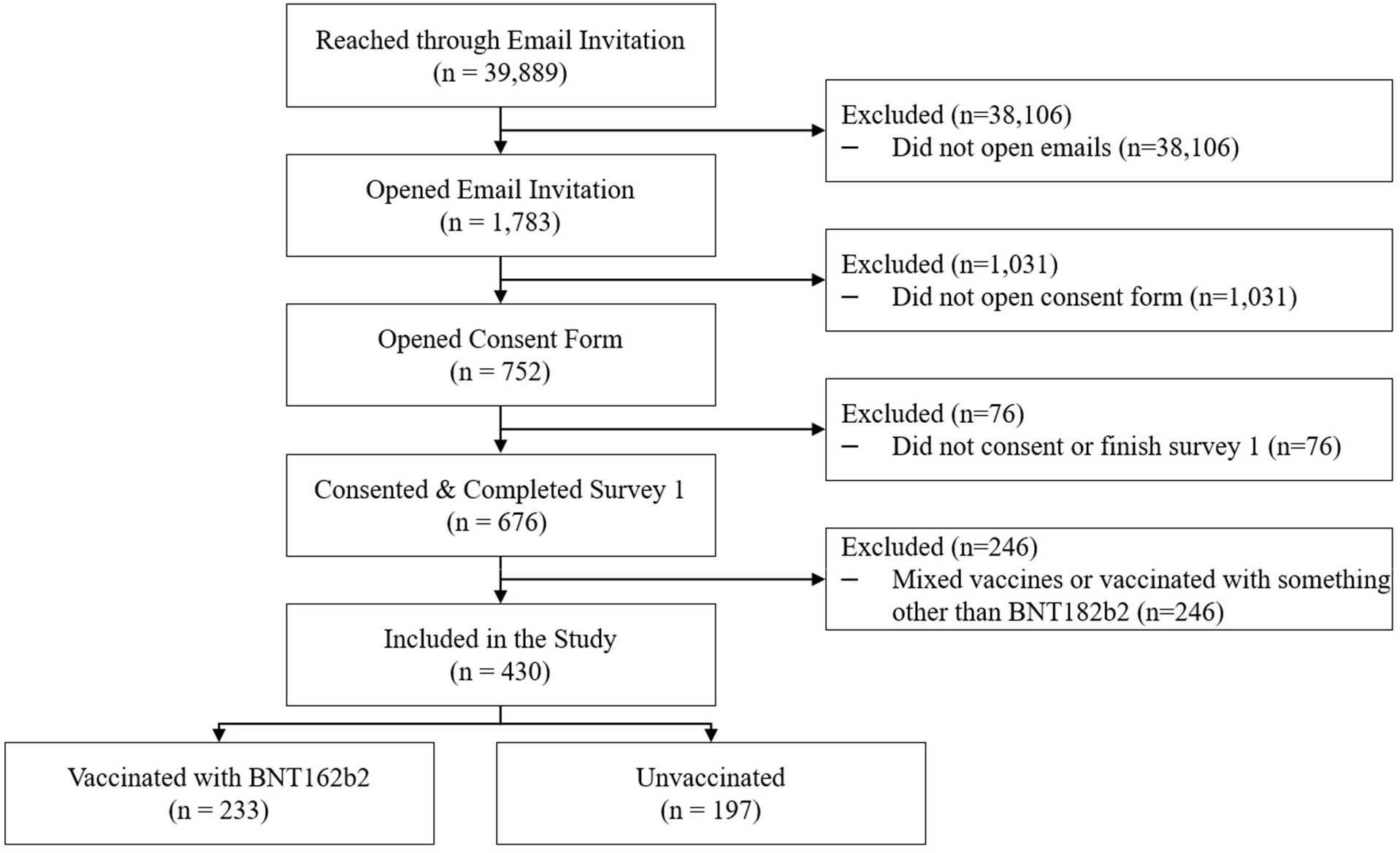
Study flow diagram.

The sociodemographic characteristics of the baseline participants are shown in Table 1. Overall, the mean (SD) age was 42.4 (14.3), 76% were female, 68.6% Caucasian, 58.7% from Southern US. There were 24.2% participants who reported ≥1 comorbidities, including 4.4% with immunocompromising conditions and 39% reported a previous COVID-19 infection.

About 46% (197) were unvaccinated and 54% (233) were vaccinated with BNT162b2; of those, respectively 140 (60%) and 93 (40%) received 2 and 3 doses. Compared with unvaccinated, BNT162b2 participants were comparable with respect to gender, working and living settings, and comorbidities, slight older with mean age 43.7 vs. 40.9 (p=0.049); living in less vulnerable area with lower mean social vulnerability index (0.40 vs. 0.49, P<<0.001); and slight differences in race/ethnicity and region. In the vaccinated group, mean (SD) time since vaccination before infection was 186 (105) days.

At index date, the most reported acute symptoms were respiratory and systemic. BNT162b2 vaccinated participants reported fewer overall acute COVID-19 symptoms on average than unvaccinated participants, mean 5.1 vs. 5.6, P=0.034 (Table 1). Directionally, the proportions of all systemic and GI-related symptoms were numerically lower in the BNT162b2 cohort. Relative to unvaccinated, those vaccinated with BNT162b2 reported significantly fewer symptoms of fever (30.5% vs. 47.2% P<0.001), chills (42.9% vs. 57.4%, P=0.003), muscle or body aches (49.4% vs. 59.4%, P=0.038), and diarrhea (15.9% vs. 25.9%, P=0.010), but more congestion or runny nose (80.7% vs. 68.0%, P=0.003).

### Post-COVID-19 symptoms

At Week 4, the mean number of symptoms was statistically lower in the BNT162B2 cohort (2.5 vs. 3.7, p=0.002). The overall prevalence decreased over time too, especially fever, cough, headache, fatigue, diarrhea, muscle pain; however, ∼70% of participants still reported at least 1 post-COVID-19 symptom. Directionally, the proportions of all symptoms were numerically lower in the BNT162b2 cohort. Symptoms of worsening after physical or mental activities (10.3% vs. 20.6%), general pain/discomfort (11.4% vs. 19.4%), change in smell or taste (10.9% vs. 20.6%), headache (16.0% vs. 25.2%), sleep problems (20.0% vs. 29.7%), mood changes (7.4% vs. 14.8%), memory loss (6.3% vs. 17.4%) and diarrhea (3.4% vs. 11.0%) were statistically significant (P < 0.05) (Table 2).

**Table 2.** Post-COVID-19 Symptoms at Week 4

| Symptom | All | BNT162b2 | Unvaccinated | P-value <sup>a</sup> |
| --- | --- | --- | --- | --- |
| General symptoms |  |  |  |  |
| Tiredness or fatigue | 136 (41.2%) | 71 (40.6%) | 65 (41.9%) | 0.802 |
| Symptoms that get worse after physical or mental activities | 50 (15.2%) | 18 (10.3%) | 32 (20.6%) | 0.009 |
| Fever | 1 (0.3%) | 0 (0.0%) | 1 (0.7%) | 0.470 |
| General pain/discomfort | 50 (15.2%) | 20 (11.4%) | 30 (19.4%) | 0.045 |
| Respiratory and cardiac |  |  |  |  |
| Difficulty breathing or shortness of breath | 58 (17.6%) | 26 (14.9%) | 32 (20.6%) | 0.168 |
| Cough | 86 (26.1%) | 40 (22.9%) | 46 (29.7%) | 0.159 |
| Chest or stomach pain | 32 (9.7%) | 14 (8.0%) | 18 (11.6%) | 0.268 |
| Fast-beating or pounding heart (also known as heart palpitations) | 38 (11.5%) | 17 (9.7%) | 21 (13.5%) | 0.276 |
| Neurologic |  |  |  |  |
| Change in smell or taste | 51 (15.5%) | 19 (10.9%) | 32 (20.6%) | 0.014 |
| Headache | 67 (20.3%) | 28 (16.0%) | 39 (25.2%) | 0.039 |
| Dizziness on standing (lightheadedness) | 45 (13.6%) | 20 (11.4%) | 25 (16.1%) | 0.214 |
| Difficulty thinking or concentrating (sometimes referred to as “brain fog”) | 86 (26.1%) | 43 (24.6%) | 43 (27.7%) | 0.513 |
| Pins-and-needles feeling | 24 (7.3%) | 10 (5.7%) | 14 (9.0%) | 0.247 |
| Sleep problems | 81 (24.5%) | 35 (20.0%) | 46 (29.7%) | 0.042 |
| Mood changes | 36 (10.9%) | 13 (7.4%) | 23 (14.8%) | 0.031 |
| Memory loss | 38 (11.5%) | 11 (6.3%) | 27 (17.4%) | 0.002 |
| Other |  |  |  |  |
| Diarrhea | 23 (7.0%) | 6 (3.4%) | 17 (11.0%) | 0.007 |
| Joint or muscle pain | 67 (20.3%) | 29 (16.6%) | 38 (24.5%) | 0.073 |
| Rash | 11 (3.3%) | 3 (1.7%) | 8 (5.2%) | 0.082 |
| Changes in period cycles | 28 (11.5%) | 12 (9.4%) | 16 (13.8%) | 0.280 |
| Number of post-COVID-19 symptoms, Mean (SD) | 3.1 (3.6) | 2.5 (3.0) | 3.7 (4.1) | 0.002 |
| 0 | 100 (30.3%) | 54 (30.9%) | 46 (29.7%) | 0.001 |
| 1-2 | 100 (30.3%) | 59 (33.7%) | 41 (26.5%) |  |
| 3-5 | 61 (18.5%) | 40 (22.9%) | 21 (13.5%) |  |
| 6-8 | 32 (9.7%) | 9 (5.1%) | 23 (14.8%) |  |
| ≥9 | 37 (11.2%) | 13 (7.4%) | 24 (15.5%) |  |

<sup>a</sup> P-values of t-test for number of symptoms, chi-square or Fisher’s exact tests when any one cell has an expected frequency less than 5 for individual symptoms and number of symptom category comparing the BNT162b2 cohort and the unvaccinated cohort.

### Health-Related Quality of life

#### Utility Index scores

Mean pre-COVID-19 baseline UIs did not differ between the BNT162b2 and unvaccinated cohorts, respectively 0.924 and 0.918 (P=0.547). COVID-19 infection had a detrimental effect on the HRQoL of participants, especially during the acute episode (Day 3). In both the BNT162b2 and the unvaccinated cohorts, UIs were lower at Day 3 and Week 4 relative to pre-COVID-19. While UIs improvement was observed over time, the UI did not return to pre-COVID levels at Week 4 (Table 3).

**Table 3.** Summary of EQ-5D-5L and WPAI-GH Scores^a^ and Their Changes from Baseline by Assessment Time

|  | BNT162b2 Cohort |  |  |  |  |  | Unvaccinated Cohort |  |  |  |  |  | Difference in Change from Baseline Between Cohorts |  |  |
| --- | --- | --- | --- | --- | --- | --- | --- | --- | --- | --- | --- | --- | --- | --- | --- |
|  | Score |  | Change from Baseline <sup>b</sup> |  |  |  | Score |  | Change from Baseline |  |  |  |  |  |  |
|  | n | Mean (SD) | n | Mean (SD) | P-value <sup>c</sup> | ES<sub>w</sub><sup>d</sup> | n | Mean (SD) | n | Mean (SD) | P-value <sup>c</sup> | ES<sub>w</sub><sup>d</sup> | Mean (SD) | P-value <sup>c</sup> | ES<sub>b</sub><sup>f</sup> |
| EQ-5D-5L |  |  |  |  |  |  |  |  |  |  |  |  |  |  |  |
| Visual analogue scale (VAS) |  |  |  |  |  |  |  |  |  |  |  |  |  |  |  |
| Baseline <sup>g</sup> | 233 | 86.9 (10.7) |  |  |  |  | 197 | 87.8 (11.0) |  |  |  |  |  | 0.414 <sup>h</sup> |  |
| Day 3 | 233 | 73.9 (15.6) | 233 | -13.0 (12.5) | <0.001 | -1.04 | 194 | 71.8 (19.6) | 194 | -16.1 (17.1) | <0.001 | -0.94 | 3.1 (14.8) | 0.033 | 0.21 |
| Week 4 | 171 | 82.9 (13.8) | 171 | -3.7 (10.7) | <0.001 | -0.35 | 151 | 80.9 (15.7) | 151 | -7.2 (13.7) | <0.001 | -0.53 | 3.5 (12.2) | 0.011 | 0.28 |
| Utility Index (US weight) |  |  |  |  |  |  |  |  |  |  |  |  |  |  |  |
| Baseline<sup>e</sup> | 233 | 0.924 (0.117) |  |  |  |  | 197 | 0.918 (0.117) |  |  |  |  |  | 0.547 <sup>h</sup> |  |
| Day 3 | 233 | 0.820 (0.193) | 233 | -0.105 (0.159) | <0.001 | -0.66 | 197 | 0.762 (0.252) | 197 | -0.155 (0.228) | <0.001 | -0.68 | 0.050 (0.194) | 0.007 | 0.26 |
| Week 4 | 176 | 0.882 (0.145) | 176 | -0.039 (0.120) | <0.001 | -0.33 | 155 | 0.838 (0.197) | 155 | -0.074 (0.156) | <0.001 | -0.47 | 0.035 (0.138) | 0.023 | 0.25 |
| WPAI-GH |  |  |  |  |  |  |  |  |  |  |  |  |  |  |  |
| Absenteeism |  |  |  |  |  |  |  |  |  |  |  |  |  |  |  |
| Baseline<sup>e</sup> | 153 | 2.8 (11.8) |  |  |  |  | 129 | 11.7 (26.4) |  |  |  |  |  | <0.001 <sup>h</sup> |  |
| Week 1 | 153 | 44.9 (38.3) | 147 | 42.5 (38.8) | <0.001 | 1.09 | 129 | 66.7 (37.0) | 127 | 55.5 (39.2) | <0.001 | 1.42 | -13.0 (39.0) | 0.006 | -0.33 |
| Week 4 | 128 | 4.6 (17.2) | 122 | 1.4 (20.1) | 0.460 | 0.07 | 106 | 3.3 (12.2) | 102 | -7.7 (28.1) | 0.007 | -0.27 | 9.0 (24.1) | 0.006 | 0.37 |
| Presenteeism |  |  |  |  |  |  |  |  |  |  |  |  |  |  |  |
| Baseline<sup>e</sup> | 153 | 9.5 (18.5) |  |  |  |  | 124 | 9.4 (20.4) |  |  |  |  |  | 0.958 <sup>h</sup> |  |
| Week 1 | 123 | 41.7 (27.4) | 119 | 33.0 (29.1) | <0.001 | 1.13 | 82 | 47.8 (34.2) | 81 | 38.2 (35.9) | <0.001 | 1.06 | -5.1 (32.1) | 0.268 | -0.16 |
| Week 4 | 125 | 11.7 (18.8) | 119 | 1.4 (24.5) | 0.527 | 0.06 | 105 | 18.7 (23.4) | 96 | 10.7 (24.1) | <0.001 | 0.45 | -9.3 (24.3) | 0.006 | -0.38 |
| Work productivity loss |  |  |  |  |  |  |  |  |  |  |  |  |  |  |  |
| Baseline<sup>e</sup> | 152 | 10.5 (20.0) |  |  |  |  | 123 | 14.2 (24.7) |  |  |  |  |  | 0.175 <sup>h</sup> |  |
| Week 1 | 123 | 56.6 (31.5) | 118 | 46.7 (34.5) | <0.001 | 1.35 | 82 | 66.9 (32.4) | 80 | 53.2 (35.3) | <0.001 | 1.51 | -6.4 (34.9) | 0.203 | -0.18 |
| Week 4 | 125 | 12.9 (20.7) | 118 | 1.4 (27.5) | 0.589 | 0.05 | 105 | 19.8 (24.6) | 96 | 7.7 (30.6) | 0.016 | 0.25 | -6.3 (29.0) | 0.113 | -0.22 |
| Activity impairment |  |  |  |  |  |  |  |  |  |  |  |  |  |  |  |
| Baseline<sup>c</sup> | 203 | 12.5 (22.2) |  |  |  |  | 176 | 17.2 (27.2) |  |  |  |  |  | 0.065<sup>h</sup> |  |
| Week 1 | 202 | 48.6 (29.8) | 202 | 36.1 (31.9) | <0.001 | 1.13 | 176 | 54.2 (32.7) | 176 | 37.0 (37.5) | <0.001 | 0.99 | -0.9 (34.6) | 0.801 | -0.03 |
| Week 4 | 175 | 15.5 (21.7) | 175 | 2.3 (26.3) | 0.242 | 0.09 | 155 | 25.9 (28.5) | 155 | 8.8 (33.8) | 0.002 | 0.26 | -6.4 (30.1) | 0.053 | -0.21 |

<sup>a</sup> Score ranges: EQ-5D-5L VAS 0 to 100, EQ-5D-5L UI (the United States weights) -0.573 to 1; WPAI-GH (absenteeism, presenteeism, work productivity loss, and activity impairment) 0 to 100 percent.
<sup>b</sup> Baseline refers to pre-COVID-19 symptom onset.
<sup>c</sup> P-value of t-test comparing mean score changes from baseline and 0 within BNT162b2 or Unvaccinated cohorts.
<sup>d</sup> ES<sub>w</sub> refers to effect size for score changes from baseline within BNT162b2 or Unvaccinated cohorts.
<sup>e</sup> P-value of t-test comparing mean score changes from baseline between BNT162b2 and Unvaccinated cohorts.
<sup>f</sup> ES<sub>b</sub> refers to effect size for score changes from baseline between BNT162b2 and Unvaccinated cohorts.
<sup>g</sup> Prior to symptom onset (pre-COVID).
<sup>h</sup> P-values of t-tests comparing pre-COVID-19 baseline mean scores between BNT162b2 and Unvaccinated cohorts.

The BNT162B2 cohort was less impacted than the unvaccinated cohort, at both Day 3 and Week 4. After controlling for pre-COVID baseline score and other covariates, the least-square estimate UI scores at Day 3 were, respectively 0.77 and 0.84 in the unvaccinated and BNT162B2 cohorts (Table 4). Moderate ESs of, respectively, 0.64 and 0.49 were observed from baseline. At Week 4, the least-square estimate UI scores were, respectively, 0.86 and 0.90. Small-to-moderate ESs of, respectively, 0.38 and 0.13 were observed from baseline. The differences between the two groups were statistically significant (P<0.05). (Table 4) Small-to-medium ESs between cohorts were observed and were 0.36 and 0.32 for Day 3 and Week 4, respectively. (Table 4, Supplemental Figure 1 and 2)

**Table 4.** Least-Square Mean Estimate and 95% Confidence Interval for EQ-5D-5L and WPAI-GH Scores ^a^

|  | BNT162b2 Cohort |  |  |  | Unvaccinated Cohort |  |  |  | Between Cohort Difference |  |  |
| --- | --- | --- | --- | --- | --- | --- | --- | --- | --- | --- | --- |
|  | Score | Change from Baseline |  |  | Score | Change from Baseline |  |  |  |  |  |
| Variable | LSE (95% CI) | LSE (95% CI) | P-value<sup>b</sup> | ES<sub>w</sub><sup>c</sup> | LSE (95% CI) | LSE (95% CI) | P-value<sup>b</sup> | ES<sub>w</sub><sup>c</sup> | LSE (95% CI) | P-value<sup>d</sup> | ES<sub>b</sub><sup>e</sup> |
| EQ-5D-5L |  |  |  |  |  |  |  |  |  |  |  |
| Visual analogue scale (VAS) |  |  |  |  |  |  |  |  |  |  |  |
| Day 3 | 76.2 (73.3, 79.2) | -11.1 (-14.1, -8.2) | <0.001 | -0.89 | 72.6 (69.7, 75.4) | -14.8 (-17.6, -11.9) | <0.001 | -0.86 | 3.6 (0.8, 6.5) | 0.013 | 0.25 |
| Week 4 | 85.0 (82.1, 87.9) | -2.3 (-5.2, 0.6) | 0.119 | -0.22 | 81.6 (78.8, 84.4) | -5.7 (-8.5, -2.9) | <0.001 | -0.42 | 3.4 (0.6, 6.2) | 0.016 | 0.28 |
| Utility Index (US weight) |  |  |  |  |  |  |  |  |  |  |  |
| Day 3 | 0.842 (0.805, 0.879) | -0.077 (-0.115, -0.040) | <0.001 | -0.49 | 0.773 (0.736, 0.809) | -0.147 (-0.183, -0.110) | <0.001 | -0.64 | 0.069 (0.032, 0.107) | <0.001 | 0.36 |
| Week 4 | 0.903 (0.868, 0.938) | -0.016 (-0.051, 0.019) | 0.369 | -0.13 | 0.859 (0.826, 0.892) | -0.060 (-0.093, -0.027) | <0.001 | -0.38 | 0.044 (0.013, 0.075) | 0.005 | 0.32 |
| WPAI-GH |  |  |  |  |  |  |  |  |  |  |  |
| Absenteeism |  |  |  |  |  |  |  |  |  |  |  |
| Week 1 | 45.6 (38.0, 53.1) | 39.1 (31.6, 46.7) | <0.001 | 1.01 | 65.0 (57.5, 72.5) | 58.6 (51.1, 66.0) | <0.001 | 1.49 | -19.4 (-28.3, -10.6) | <0.001 | -0.50 |
| Week 4 | 5.3 (0.0, 10.7) | -1.1 (-6.5, 4.2) | 0.676 | -0.06 | 2.2 (0.0, 7.1) <sup>f</sup> | -4.3 (-9.2, 0.6) | 0.086 | -0.15 | 3.2 (-1.1, 7.4) | 0.145 | 0.13 |
| Presenteeism |  |  |  |  |  |  |  |  |  |  |  |
| Week 1 | 38.4 (30.3, 46.5) | 29.0 (20.8, 37.1) | <0.001 | 1.00 | 46.8 (38.6, 55.1) | 37.4 (29.1, 45.6) | <0.001 | 1.04 | -8.4 (-16.7, -0.1) | 0.047 | -0.26 |
| Week 4 | 6.8 (0, 13.9) <sup>f</sup> | -2.7 (-9.7, 4.4) | 0.458 | -0.11 | 16.0 (9.4, 22.5) | 6.5 (0.0, 13.0) | 0.050 | 0.27 | -9.2 (-14.7, -3.6) | 0.001 | -0.38 |
| Work productivity loss |  |  |  |  |  |  |  |  |  |  |  |
| Week 1 | 53.8 (45.1, 62.4) | 42.4 (33.8, 51.0) | <0.001 | 1.23 | 65.0 (56.2, 73.7) | 53.6 (44.8, 62.4) | <0.001 | 1.52 | -11.2 (-19.9, -2.5) | 0.012 | -0.32 |
| Week 4 | 8.6 (0.9, 16.2) | -2.8 (-10.4, 4.9) | 0.476 | -0.10 | 17.0 (9.9, 24.0) | 5.7 (-1.4, 12.7) | 0.116 | 0.18 | -8.4 (-14.5, -2.3) | 0.007 | -0.29 |
| Activity impairment |  |  |  |  |  |  |  |  |  |  |  |
| Week 1 | 43.9 (37.7, 50.1) | 29.0 (22.8, 35.3) | <0.001 | 0.91 | 50.2 (44.2, 56.2) | 35.4 (29.3, 41.4) | <0.001 | 0.94 | -6.3 (-12.4, -0.2) | 0.044 | -0.18 |
| Week 4 | 11.0 (5.1, 16.9) | -3.9 (-9.7, 2.0) | 0.194 | -0.15 | 21.3 (15.7, 26.9) | 6.4 (0.8, 12.0) | 0.025 | 0.19 | -10.3 (-15.6, -5.0) | <0.001 | -0.34 |

Abbreviations: LSE = Least-Square Mean Estimate; CI = Confidence Interval.
<sup>a</sup> Multivariate models include variables for time, vaccination status and interaction of time by vaccination status, as well as covariates of participant pre-COVID-19 symptom onset score, sociodemographic characteristics (age, sex, regions, social vulnerability, race/ethnicity, high risk occupations), previously tested positive for COVID-19, severity of acute illness (number of symptoms reported on index date), and immunocompromised status. Parameter estimates are presented in Supplemental Table 3.
<sup>b</sup> P-value refers to the comparison of least-square mean estimates score changes from baseline and 0 within BNT162b2 or Unvaccinated cohorts
<sup>c</sup> ES<sub>w</sub>, within-cohort effect size, was calculated as the least square estimate of mean change from divided by the observed standard deviation of change scores from baseline to follow-up.
<sup>d</sup> ES<sub>b</sub>, between-cohort effect size, was calculated as the difference in least square estimates of mean changes from baseline between cohorts, divided by the observed pooled standard deviation of change scores
<sup>e</sup> P-value refers to the difference in least-square mean estimates between BNT162b2 and Unvaccinated cohorts.
<sup>f</sup> Lower limit of 95% CI was truncated from -2.7 to 0 for absenteeism and -0.3 to 0 for presenteeism at Week 4 because the valid range is 0 to 100.

#### EQ-VAS

The pattern of EQ-VAS scores was similar to that observed for UIs. Mean pre-COVID-19 baseline EQ-VAS were similar for the BNT162b2 and unvaccinated cohorts, respectively 86.9 and 87.8 (P=0.414) (Table 3). Similar to the UIs, the pre-COVID EQ-VAS were rated relatively high by the participants, indicating a generally healthy cohort. The least-square estimate EQ-VAS scores for the BNT162b2 and unvaccinated cohorts were, respectively, 76.2 and 72.6 at Day 3 and 85.0 and 81.6 at Week 4. After controlling for pre-COVID-19 baseline score and other covariates, the least-square estimates of change from pre-COVID-19 baseline in EQ VAS for the BNT162B2 and the unvaccinated cohort were −11.1 and −14.8, respectively on Day 3, and −2.3 and −5.7, respectively at Week 4. COVID-19 had a large adverse impact on EQ-VAS with an ES of −0.89 for BNT162B2 cohort and −0.86 for Unvaccinated cohort on Day 3, and small ES (−0.22) for BNT162B2 cohort and approaching medium ES (−0.42) for Unvaccinated cohort at Week 4. BNT162B2 cohort was associated with 3.6 (P=0.013) on Day 3 and 3.4 (P=0.016) at Week 4 less drop in EQ VAS than the Unvaccinated cohort. The ESs between cohorts were small yet relevant, being 0.25 and 0.28 for Day 3 and Week 4, respectively (Table 4, Supplemental Figure 1 and 2).

#### EQ-5D-5L dimensions

The health status of the study participants according to the dimensions of EQ-5D-5L is reported in Figure 3 and Supplemental Table 2. In both groups, at Day 3, over half of the cohort reported problems in usual activities, pain/discomfort and anxiety/depression, while the vast majority reported no or slight problems in mobility and self-care. At Week 4, the vast majority continued to report no or slight problems with mobility, self-care, as well as for usual activities; most reported no, slight or moderate problems with pain/discomfort and anxiety/depression. BNT162b2 cohort had lower mean responses across all 5 domains at both Day 3 and Week 4 relative to unvaccinated.

**Figure 3.**
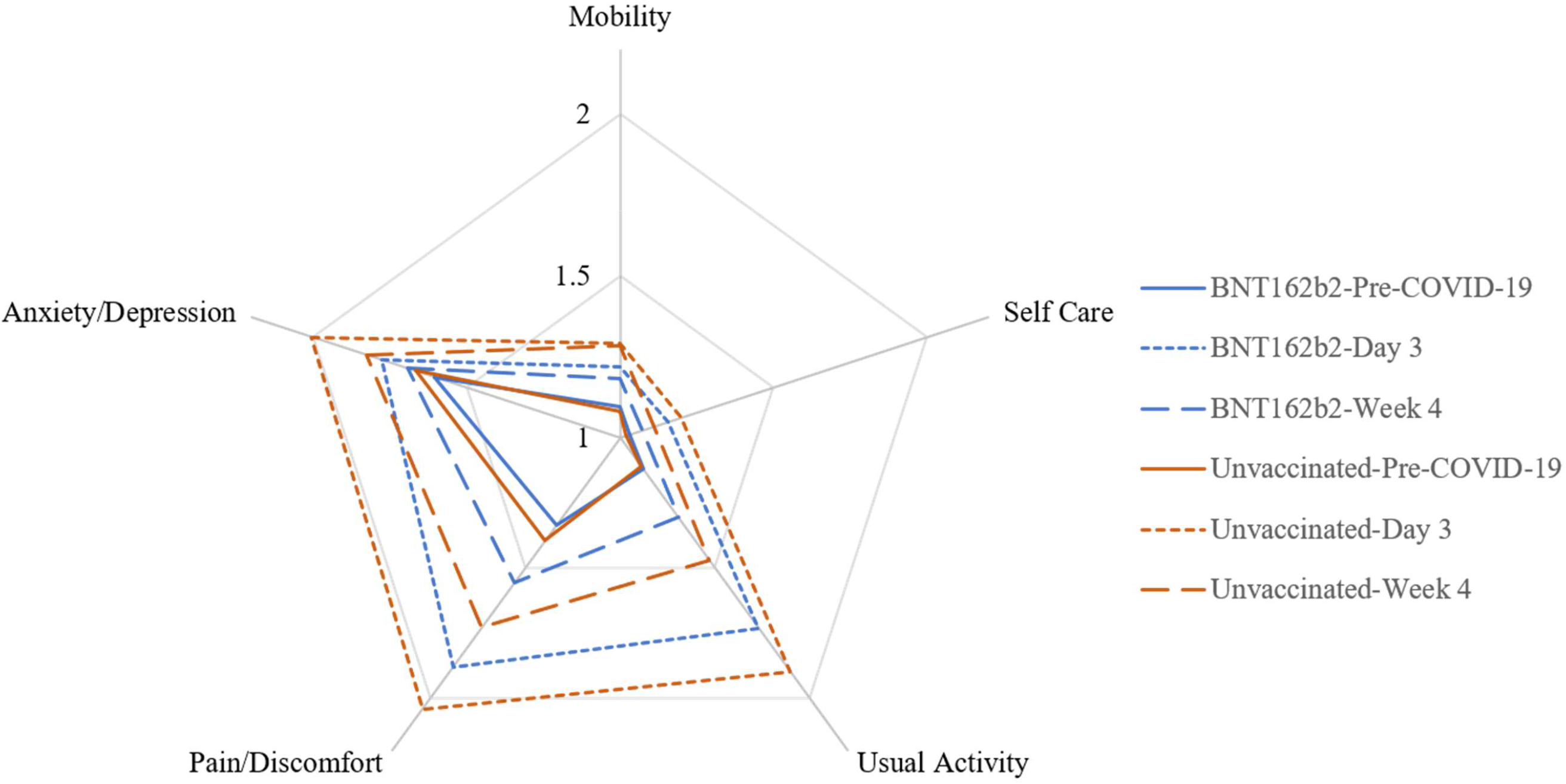
Mean Responses of EQ-5D-5L Dimensions by Timepoint Mean dimension scores range from 1 for no problem to 5 for extreme / unable. The blue and red solid lines indicate that vaccinated and unvaccinated were similar at the pre-COVID baseline. At Day 3 and Week 4 post-index date, vaccinated cohort was less impacted (lower scores) than unvaccinated by COVID on anxiety/depression, pain/discomfort, and usual activities (dotted lines for Day 3, dashed lines for Week 4).

### Work Productivity and Activity Impairment

Approximately 65% of participants reported being currently employed at baseline (155 in the BNT162b2 cohort and 129 unvaccinated), and were eligible to complete the absenteeism, presenteeism and work-productivity loss questions. At Week 1, COVID-19 had a large impact on all four WPAI-GH domains for both the unvaccinated and BNT162b2cohort. The mean time loss due to absenteeism was, respectively, 65.0% and 45.6%; the mean time loss due to presenteeism was, respectively, 46.8% and 38.4%; the mean time of work productivity loss was 65.0% and 53.8%, and the mean time of activity impairment was 50.2% and 43.9%. All within-cohort Ess were > 0.8, which are considered large effects (Table 3). After controlling for pre-COVID-19 baseline score, and other covariates, the BNT162b2 cohort was associated with less worsening in WPAI-GH scores. Small-to-medium ESs were observed for work-related scores (absenteeism −0.50, presenteeism −0.26, and work productivity loss −0.32) between the BNT162b2 cohort and the unvaccinated cohort (Table 4). At Week 4, the mean time loss dropped across all four domains. The time loss due to absenteeism dropped substantially; the change from baseline in absenteeism was not found to be statistically significant between the BNT162b2 cohort and the unvaccinated cohort. Small-to-medium ESs were observed for presenteeism (−0.38) work productivity loss (−0.29), and activity impairment (−0.34) between the BNT162b2 cohort and the unvaccinated cohort (Table 4, Supplemental Figure 3 and 4).

## DISCUSSION

The impacts of SARS-CoV-2 infection go beyond its clinical outcomes.

We found that mild acute infection can negatively impact the humanistic outcomes for up to four weeks post infection. Shortly after infection, the UI and EQ-VAS HRQoL scores dropped from pre-COVID, and over half of the study population reported problems in usual activities, pain/discomfort and anxiety/depression. At Week 1, the work productivity and activity impairment time loss were over 50%. At Week 4, both the HRQoL and WPAI scores improved, although they did not return to pre-COVID levels. Individuals vaccinated with BNT162b2 were less impacted and recovered faster than unvaccinated individuals. Multivariable analyses showed that BNT162b2 was significantly associated with higher EQ-VAS and UI scores, less symptoms and better WPAI scores, except for absenteeism at Week 4.

There is limited evidence measuring the health-related wellbeing of non-hospitalized individuals affected by COVID-19 [1–4]. To our knowledge, this is the first report measuring the impact of COVID-19 on the HRQoL and WPAI among a national sample of outpatients in the United States. In contrast to our study, previous research that used EQ-5D scales to measure COVID-19 impact on the HRQoL reported mean UI scores ranging from 0.61 to 0.86 depending on the hospitalization treatment and time since discharge [1, 2]. The EQ-VAS scores ranged from 50.7 to 70.3 [1, 2]. In our study, the HRQoL scores at Day 3 and Week 4 among outpatients with mild disease are higher than those, likely due to the different study populations and periods. In a small US study assessing the impact of COVID-19 on WPAI ∼4 months post-infection among subjects enrolled in clinical trials before the introduction of vaccines, 46% of the non-hospitalized patients reported health-related impairment in daily activities [24]. Among the employed, 11.5% missed work and 38.9% reported impairment at work due to health. In our study, all the WPAI scores among unvaccinated at Week 1 are higher, and those at Week 4 similar or lower than those, likely due to the different cut-off, study populations, periods and design.

HRQoL and WPAI studies are scarce in COVID-19 related vaccination research. To our knowledge, this is the first report on the effects of BNT162b2 on these patient-centric outcomes. These results indicate an additive benefit beyond vaccine effectiveness that should be explored further.

Strengths of this study include the nationally representative real-world source population of mildly symptomatic outpatients, the prospective collection of the primary outcomes via validated instruments, and the representativeness of the employed population for work productivity analyses (∼65% of the cohort). The age distribution of study participants was comparable with the non-enrolled tested population (p=0.076 for mean age, Supplemental Table 1) and with CDC research in non-hospitalized adults: 91.6% were between 18 and 64 and 8.4% were 65 or older, which was quite similar to Hernandez-Romieu [25], 92.7% and 7.3%, respectively. In the vaccinated group, the time between vaccination and breakthrough infection (mean: ∼6 months) was consistent with literature on vaccine-induced duration of protection [5].

The study findings are subject to limitations. All the data analyzed was self-reported and may be subject to error, missingness, recall bias, social desirability bias, and selection bias associated with survey drop-out. Out of 430 participants completing Day 3 survey, 12% (51/430) missed Week 1 and 23% (99/430) missed Week 4 survey. The drop in responses may partly be the result of responders’ fatigue, and/or recovered cases not returning to follow-up surveys. Female and older age (≥30 years) were found to be more likely to miss follow-up surveys. However, after taking into account several variables, the model predicting missingness indicated that missingness was not associated with vaccination status, nor with HRQoL in terms of EQ VAS on Day 3 and its change from pre-COVID-19 baseline (Supplemental Table 4).

The study population differed from non-enrolled tested outpatients in being predominantly female, white, with a higher vaccination uptake, and slightly more comorbidities (Supplemental Table 1). The female over-representation is in line with prior research indicating that women are more likely to contribute to health research surveys [2]. In our study, ∼24% of participants reported at least one comorbidity, in contrast to 20% among the non-enrolled tested and ∼35% in Hernandez-Romieu (2021) [25]. Various models were fit to account for potential effects due to sociodemographic factors and comorbidities. These adjusted effect sizes between BNT162b2 and unvaccinated cohorts were similar and consistent with those calculated from observed data (unadjusted effect sizes).

The pre-COVID baseline scores were slightly higher than US population norms. The mean EQ-VAS and UI were 86.9 and 0.924 for the BNT162b2 cohort, and 87.8 and 0.918 for the unvaccinated cohort, respectively. Cha et al (2019) reported a mean EQ-VAS of 84.6 for the U.S. general population [12]. Jiang et al (2021) reported the US population norm of EQ-VAS as 80.4 and the mean EQ-5D-5L UI as 0.851 [26]. The healthy pre-infection status of the study population and the potential for retrospective recall bias may partially explain the differences. A modified EQ-5D-5L questionnaire was used to retrospectively measure pre-COVID-19 baseline data; despite literature suggesting concordance between prospective and retrospective measurements of EQ-5D-5L [27, 28], and a good correlation between assessment of baseline before the index date and recall assessment of the baseline after index date, there is currently no information regarding the recall application of the EQ-5D-5L for COVID-related studies. The pre-COVID-19 values for abseenteism and presenteeism were 3.1% and 9.5% in the BNT162b2 cohort, and were generally in line with Tundia et al (2015) [29], whom reported 4% absenteeism and 10% presenteeism for the US general population. The reported values were slightly higher among unvaccinated, respectively 11.7% and 9.4%.

There is currently no standard definition of minimal clinically important difference of PROs in COVID-19 research. We used effect size (ES) of Cohen’s d to quantify the magnitude of score change from baseline within the BNT162b2 vaccinated cohort and the unvaccinated cohort, as well the difference between these two cohorts [20]. An effect size ES of 0 between groups indicates that the average (typical) treated vaccinated person has a score that is no different from the typical control person; equivalently, scores of the typical treated person are more favorable than 50% of the individual scores in the control group, meaning no incremental benefit (a coin toss as to which intervention is better). If the vaccinated cohort is presumed more effective than the unvaccinated cohort, effect sizes thresholds of 0.2, 0.3, 0.5, and 0.8 (in absolute value) indicate that, based on the standardized normal distribution, the score of the typical person in the vaccinated cohort is more favorable than 58% (8% incremental benefit), 62% (12% incremental benefit), 69% (19% incremental benefit), and 70% (29% increment benefit) of the scores from individuals in the unvaccinated cohort. For example, from baseline to Week 1, the increase in the absenteeism WPAI score of the typical person in the vaccinated cohort was less (more favorable) than the corresponding change in 69% of individuals in the unvaccinated group (effect size = −0.5). Depending on the type of outcome, the same type of effect size interpretation for between cohorts can be given within cohort. For instance, from baseline to Week 4, the quality of life EQ-VAS score of the typical person in the vaccine cohort at Week 4 was less (worse) than 59% of the individual scores in the vaccine cohort at baseline (effect size = −0.22); the quality of life EQ-VAS score of the typical person in the unvaccinated cohort at Week 4 was less (worse) than 66% of the individual scores in the unvaccinated cohort at baseline (effect size = −0.42).

The study did not assess the impact on pediatrics, caregivers, long-term outcomes (e.g., “Long COVID”), and the data was collected during Omicron predominance in the US. Therefore, these findings may not be generalizable to prior or future variants, other countries, time periods and populations that were excluded. COVID-19 sequalae can affect a substantial portion of patients, with long-term consequences for their health, continuity of care and ability to work [1, 2]. Persistent symptoms and work impairment were reported ∼4 months after infection among non-hospitalized US patients enrolled in clinical trials [24]. Continued follow-up studies covering longer time periods may inform whether the protection provided by COVID-19 vaccination extends beyond the acute phase. Only generic PROMs were used in this study; COVID-19 disease-specific instruments are under development [30, 31], warranting research on their implementation. The PROMs omitted questions on vaccine adverse events. The mean 6-month interval between vaccination and breakthrough infection and a medical review ruled out cases of residual symptoms from vaccination. Research on the impact of vaccine adverse events on HRQoL is warranted.

Lastly, the study adopted an observational design, which is limited in establishing causal relationships. Future studies using different data collection methods could corroborate the study findings, including those with more rigorous study design.

Consistent with literature, our study found that COVID-19 is detrimental to mildly symptomatic COVID-19 patients. The findings provide a meaningful contribution suggesting that the ability of BNT162b2 to reduce adverse outcomes from COVID-19 disease can translate to lessening the broad impact of COVID-19 and improvements in quality of life, work productivity and activity.

## CONCLUSION

This study found that mild COVID-19 infection at a time of Omicron predominance adversely impacted the HRQoL, daily activity and work productivity of patients. This detrimental effect improved over time, although it persisted for at least one month post infection. Compared with unvaccinated, those vaccinated with BNT162b2 were less impacted and recovered faster. These findings advance research on COVID-19 associated humanistic outcomes and the potential effect of BNT162b2 in lessening the loss of HRQoL, daily activity and work productivity due to COVID-19. The results can inform the estimation of quality-adjusted life years and indirect cost savings in health economic studies.

## Data Availability

Aggregated data that support the findings of this study are available upon reasonable request from the corresponding author MDF, subject to review. These data are not publicly available due to them containing information that could compromise research participant privacy/consent.

## List of Abbreviations

CDC: Centers for Disease Control and Prevention;
CI: Confidence interval;
EQ-5D-5L: EuroQoL Group 5 dimension and 5 level questionnaire;
ES: Effect size;
GH: General Health;
HRQoL: Health Related Quality of Life;
MMRM: Mixed models for repeated measures;
SD: standard deviation;
SE: standard error;
STROBE: the Strengthening the Reporting of Observational Studies in Epidemiology;
UI: utility index;
VAS: Visual analogue scale;
WPAI: Work Productivity and Impairment.

## DECLARATIONS

### Ethics Approval and consent to participate

This study was approved by the Sterling IRB, Protocol #C4591034. Participation in the study was voluntary and anonymous. Consent was obtained electronically via the CVS Health E-Consent platform. Participants were informed of their right to refuse or withdraw from the study at any time. Participants were compensated for their time.

### Consent for publication

All authors have given their approval for this manuscript version to be published.

### Authors’ contributions

All named authors meet the International Committee of Medical Journal Editors (ICMJE) criteria for authorship for this article. All authors contributed to study conception and design, data acquisition, analysis, and interpretation, drafting and revising of the manuscript.

## Acknowledgements

The authors acknowledge Alejandro Cane, Deepa Malhotra and Nancy Gifford (Pfizer employees), Joseph Ferenchick, Shiyu Lin and Shawn Edmonds (CVS Health employees) for specific contributions to this research project. Editorial support was provided by Laura Anatale-Tardiff and Leena Samuel at CVS Health and was funded by Pfizer.

---

**Supplemental Table 1.** Patient Characteristics by Enrollment Status

| Patient characteristics | Enrolled<br>N=676 | Not-Enrolled<br>N=39,213 | P value |
| --- | --- | --- | --- |
| Vaccinated |  |  | <0.001 |
| Yes | 465 (68.8%) | 22,125 (56.4%) |  |
| No | 211 (31.2%) | 17,087 (43.6%) |  |
| Missing |  |  |  |
| Age, years |  |  |  |
| Mean, SD | 43.2 (14.7) | 42.1 (15.5) | 0.076 |
| Median, Q1, Q3 | 41 (31--55) | 40 (29--54) | 0.010 |
| Age Category |  |  |  |
| 18-29 | 134 (19.8%) | 10,095 (25.7%) | 0.005 |
| 30-49 | 317 (46.9%) | 16,706 (42.6%) |  |
| 50-64 | 156 (23.1%) | 8,561 (21.8%) |  |
| ≥65 | 69 (10.2%) | 3,850 (9.8%) |  |
| Gender, n (%) |  |  | <0.001 |
| Male | 181 (26.8%) | 17,843 (45.5%) |  |
| Female | 495 (73.2%) | 21,370 (54.5%) |  |
| Race / Ethnicity, n (%) |  |  | <0.001 |
| White or Caucasian (non-Hispanic or Latino) | 486 (71.9%) | 22,647 (57.8%) |  |
| Black or African American | 32 (4.7%) | 4,162 (10.6%) |  |
| Hispanic or Latino | 85 (12.6%) | 6,878 (17.5%) |  |
| Asian | 35 (5.2%) | 2,551 (6.5%) |  |
| Patient Refused | 16 (2.4%) | 1,535 (3.9%) |  |
| Other | 22 (3.3%) | 1,439 (3.7%) |  |
| US Geographic Region |  |  | 0.258 |
| Northeast | 92 (13.6%) | 4,387 (11.2%) |  |
| South | 402 (59.5%) | 24,305 (62.0%) |  |
| Midwest | 118 (17.5%) | 6,414 (16.4%) |  |
| West | 64 (9.5%) | 4,105 (10.5%) |  |
| Number of acute COVID-19 symptoms, Mean (SD) | 5.2 (2.5) | 5.1 (2.5) | 0.732 |
| Number of comorbidities, Mean (SD) | 0.3 (0.7) | 0.3 (0.6) | 0.021 |
| ≥1 comorbidity | 164 (24.3%) | 7,677 (19.6%) | 0.002 |

**Supplemental Table 2.** Summary of EQ-5D-5L Dimensions

| Dimension | All | BNT162b2 | Unvaccinated | P-value <sup>a</sup> |
| --- | --- | --- | --- | --- |
| Pre-COVID-19 Baseline |  |  |  |  |
| Mobility |  |  |  | 0.841 |
| No problems | 396 (92.1%) | 213 (91.4%) | 183 (92.9%) |  |
| Slight problems | 30 (7.0%) | 18 (7.7%) | 12 (6.1%) |  |
| Moderate problems | 4 (0.9%) | 2 (0.9%) | 2 (1.0%) |  |
| Severe problems | 0 (0.0%) | 0 (0.0%) | 0 (0.0%) |  |
| Unable | 0 (0.0%) | 0 (0.0%) | 0 (0.0%) |  |
| Missing | 0 (0.0%) | 0 (0.0%) | 0 (0.0%) |  |
| Self-Care |  |  |  | 0.560 |
| No problems | 419 (97.4%) | 226 (97.0%) | 193 (98.0%) |  |
| Slight problems | 11 (2.6%) | 7 (3.0%) | 4 (2.0%) |  |
| Moderate problems | 0 (0.0%) | 0 (0.0%) | 0 (0.0%) |  |
| Severe problems | 0 (0.0%) | 0 (0.0%) | 0 (0.0%) |  |
| Unable | 0 (0.0%) | 0 (0.0%) | 0 (0.0%) |  |
| Missing | 0 (0.0%) | 0 (0.0%) | 0 (0.0%) |  |
| Usual Activities |  |  |  | 0.904 |
| No problems | 394 (91.6%) | 214 (91.8%) | 180 (91.4%) |  |
| Slight problems | 23 (5.4%) | 11 (4.7%) | 12 (6.1%) |  |
| Moderate problems | 12 (2.8%) | 7 (3.0%) | 5 (2.5%) |  |
| Severe problems | 1 (0.2%) | 1 (0.4%) | 0 (0.0%) |  |
| Unable | 0 (0.0%) | 0 (0.0%) | 0 (0.0%) |  |
| Missing | 0 (0.0%) | 0 (0.0%) | 0 (0.0%) |  |
| Pain / Discomfort |  |  |  | 0.614 |
| No | 307 (71.4%) | 172 (73.8%) | 135 (68.5%) |  |
| Slight | 93 (21.6%) | 46 (19.7%) | 47 (23.9%) |  |
| Moderate | 27 (6.3%) | 13 (5.6%) | 14 (7.1%) |  |
| Severe | 3 (0.7%) | 2 (0.9%) | 1 (0.5%) |  |
| Extreme | 0 (0.0%) | 0 (0.0%) | 0 (0.0%) |  |
| Missing | 0 (0.0%) | 0 (0.0%) | 0 (0.0%) |  |
| Anxiety / Depression |  |  |  | 0.395 |
| No | 230 (53.5%) | 124 (53.2%) | 106 (53.8%) |  |
| Slightly | 140 (32.6%) | 80 (34.3%) | 60 (30.5%) |  |
| Moderately | 49 (11.4%) | 26 (11.2%) | 23 (11.7%) |  |
| Severely | 10 (2.3%) | 3 (1.3%) | 7 (3.6%) |  |
| Extremely | 1 (0.2%) | 0 (0.0%) | 1 (0.5%) |  |
| Missing | 0 (0.0%) | 0 (0.0%) | 0 (0.0%) |  |
| Days 3 |  |  |  |  |
| Mobility |  |  |  | 0.131 |
| No problems | 343 (79.8%) | 193 (82.8%) | 150 (76.1%) |  |
| Slight problems | 68 (15.8%) | 31 (13.3%) | 37 (18.8%) |  |
| Moderate problems | 17 (4.0%) | 7 (3.0%) | 10 (5.1%) |  |
| Severe problems | 2 (0.5%) | 2 (0.9%) | 0 (0.0%) |  |
| Unable | 0 (0.0%) | 0 (0.0%) | 0 (0.0%) |  |
| Missing | 0 (0.0%) | 0 (0.0%) | 0 (0.0%) |  |
| Self-Care |  |  |  | 0.571 |
| No problems | 370 (86.0%) | 205 (88.0%) | 165 (83.8%) |  |
| Slight problems | 47 (10.9%) | 21 (9.0%) | 26 (13.2%) |  |
| Moderate problems | 11 (2.6%) | 6 (2.6%) | 5 (2.5%) |  |
| Severe problems | 2 (0.5%) | 1 (0.4%) | 1 (0.5%) |  |
| Unable | 0 (0.0%) | 0 (0.0%) | 0 (0.0%) |  |
| Missing | 0 (0.0%) | 0 (0.0%) | 0 (0.0%) |  |
| Usual Activities |  |  |  | 0.361 |
| No problems | 207 (48.1%) | 117 (50.2%) | 90 (45.7%) |  |
| Slight problems | 132 (30.7%) | 75 (32.2%) | 57 (28.9%) |  |
| Moderate problems | 64 (14.9%) | 30 (12.9%) | 34 (17.3%) |  |
| Severe problems | 21 (4.9%) | 9 (3.9%) | 12 (6.1%) |  |
| Unable | 6 (1.4%) | 2 (0.9%) | 4 (2.0%) |  |
| Missing | 0 (0.0%) | 0 (0.0%) | 0 (0.0%) |  |
| Pain / Discomfort |  |  |  | 0.080 |
| No | 145 (33.7%) | 85 (36.5%) | 60 (30.5%) |  |
| Slight | 183 (42.6%) | 99 (42.5%) | 84 (42.6%) |  |
| Moderate | 82 (19.1%) | 43 (18.5%) | 39 (19.8%) |  |
| Severe | 17 (4.0%) | 4 (1.7%) | 13 (6.6%) |  |
| Extreme | 3 (0.7%) | 2 (0.9%) | 1 (0.5%) |  |
| Missing | 0 (0.0%) | 0 (0.0%) | 0 (0.0%) |  |
| Anxiety / Depression |  |  |  | 0.047 |
| No | 190 (44.2%) | 108 (46.4%) | 82 (41.6%) |  |
| Slightly | 135 (31.4%) | 79 (33.9%) | 56 (28.4%) |  |
| Moderately | 75 (17.4%) | 37 (15.9%) | 38 (19.3%) |  |
| Severely | 26 (6.1%) | 8 (3.4%) | 18 (9.1%) |  |
| Extremely | 4 (0.9%) | 1 (0.4%) | 3 (1.5%) |  |
| Missing | 0 (0.0%) | 0 (0.0%) | 0 (0.0%) |  |
| Week 4 |  |  |  |  |
| Mobility |  |  |  | 0.278 |
| No problems | 270 (81.3%) | 149 (84.7%) | 121 (77.6%) |  |
| Slight problems | 49 (14.8%) | 22 (12.5%) | 27 (17.3%) |  |
| Moderate problems | 12 (3.6%) | 5 (2.8%) | 7 (4.5%) |  |
| Severe problems | 1 (0.3%) | 0 (0.0%) | 1 (0.6%) |  |
| Unable | 0 (0.0%) | 0 (0.0%) | 0 (0.0%) |  |
| Missing | 1 (0.0%) | 1 (0.0%) | 0 (0.0%) |  |
| Self-Care |  |  |  | 0.068 |
| No problems | 308 (93.1%) | 167 (94.9%) | 141 (91.0%) |  |
| Slight problems | 17 (5.1%) | 5 (2.8%) | 12 (7.7%) |  |
| Moderate problems | 5 (1.5%) | 4 (2.3%) | 1 (0.7%) |  |
| Severe problems | 1 (0.3%) | 0 (0.0%) | 1 (0.7%) |  |
| Unable | 0 (0.0%) | 0 (0.0%) | 0 (0.0%) |  |
| Missing | 2 (0.0%) | 1 (0.0%) | 1 (0.0%) |  |
| Usual Activities |  |  |  | 0.152 |
| No problems | 232 (70.1%) | 132 (75.0%) | 100 (64.5%) |  |
| Slight problems | 75 (22.7%) | 35 (19.9%) | 40 (25.8%) |  |
| Moderate problems | 20 (6.0%) | 8 (4.6%) | 12 (7.7%) |  |
| Severe problems | 4 (1.2%) | 1 (0.6%) | 3 (1.9%) |  |
| Unable | 0 (0.0%) | 0 (0.0%) | 0 (0.0%) |  |
| Missing | 2 (0.0%) | 1 (0.0%) | 1 (0.0%) |  |
| Pain / Discomfort |  |  |  | 0.097 |
| No | 170 (51.4%) | 95 (54.0%) | 75 (48.4%) |  |
| Slight | 118 (35.6%) | 66 (37.5%) | 52 (33.5%) |  |
| Moderate | 37 (11.2%) | 13 (7.4%) | 24 (15.5%) |  |
| Severe | 5 (1.5%) | 2 (1.1%) | 3 (1.9%) |  |
| Extreme | 1 (0.3%) | 0 (0.0%) | 1 (0.7%) |  |
| Missing | 2 (0.0%) | 1 (0.0%) | 1 (0.0%) |  |
| Anxiety / Depression |  |  |  | 0.605 |
| No | 167 (50.5%) | 92 (52.3%) | 75 (48.4%) |  |
| Slightly | 98 (29.6%) | 53 (30.1%) | 45 (29.0%) |  |
| Moderately | 51 (15.4%) | 25 (14.2%) | 26 (16.8%) |  |
| Severely | 10 (3.0%) | 5 (2.8%) | 5 (3.2%) |  |
| Extremely | 5 (1.5%) | 1 (0.6%) | 4 (2.6%) |  |
| Missing | 2 (0.0%) | 1 (0.0%) | 1 (0.0%) |  |

<sup>a</sup> P values of Freeman-Halton tests comparing BNT162b2 cohort and unvaccinated cohort, excluding category of missing.

**Supplemental Table 3.** Mixed Models for Repeated Measurements EQ-5D-5L and WPAI-GH Scores

|  | EQ VAS |  | EQ-5D-5L Utility Index<br>(U.S. weights) |  | Absenteeism |  | Presenteeism |  | Work productivity<br>loss |  | Activity impairment |  |
| --- | --- | --- | --- | --- | --- | --- | --- | --- | --- | --- | --- | --- |
|  | Coeff (SE) | P value | Coeff (SE) | P value | Coeff (SE) | P value | Coeff (SE) | P value | Coeff (SE) | P value | Coeff (SE) | P value |
| Intercept | 4.51 (5.36) | 0.401 | 0.038 (0.062) | 0.546 | 53.6 (4.9) | <0.001 | 41.4 (5.7) | <0.001 | 57.7 (6.0) | <0.001 | 41.2 (4.9) | <0.001 |
| Vaccinated |  |  |  |  |  |  |  |  |  |  |  |  |
| BNT162B2BNT162b2 | 3.65 (1.46) | 0.013 | 0.069 (0.019) | <0.001 | -19.4 (4.5) | <0.001 | -8.4 (4.2) | 0.047 | -11.2 (4.4) | 0.012 | -6.3 (3.1) | 0.044 |
| No | Reference |  | Reference |  | Reference |  | Reference |  | Reference |  | Reference |  |
| Assessment Time |  |  |  |  |  |  |  |  |  |  |  |  |
| Day 2-4 / Week 1 | Reference |  | Reference |  | Reference |  | Reference |  | Reference |  | Reference |  |
| Week 4 | 9.04 (1.23) | <0.001 | 0.086 (0.014) | <0.001 | -62.9 (3.4) | <0.001 | -30.9 (3.3) | <0.001 | -48.0 (3.6) | <0.001 | -28.9 (2.5) | <0.001 |
| Assessment Time * Vaccinated |  |  |  |  |  |  |  |  |  |  |  |  |
| Day 2-4 / Week 1 *<br>BNT162b2 | Reference |  | Reference |  | Reference |  | Reference |  | Reference |  | Reference |  |
| Week 4 * BNT162B2 | -0.25 (1.69) | 0.884 | -0.025 (0.020) | 0.209 | 22.6 (4.8) | <0.001 | -0.8 (4.4) | 0.863 | 2.8 (4.8) | 0.561 | -4.0 (3.5) | 0.257 |
| Pre-COVID-19 Baseline<br>Score | -0.22 (0.05) | <0.001 | -0.177 (0.057) | 0.002 | -0.8 (0.0) | <0.001 | -0.7 (0.1) | <0.001 | -0.8 (0.1) | <0.001 | -0.7 (0.0) | <0.001 |
| Age, years |  |  |  |  |  |  |  |  |  |  |  |  |
| 18-29 | Reference |  | Reference |  | Reference |  | Reference |  | Reference |  | Reference |  |
| 30-49 | -1.82 (1.45) | 0.211 | -0.011 (0.018) | 0.532 | 3.9 (2.4) | 0.102 | 5.6 (3.0) | 0.065 | 4.1 (3.2) | 0.204 | 7.8 (2.9) | 0.006 |
| 50-64 | -2.84 (1.74) | 0.102 | -0.048 (0.021) | 0.025 | 10.8 (2.9) | <0.001 | 6.8 (3.8) | 0.074 | 7.3 (4.1) | 0.075 | 13.7 (3.5) | <0.001 |
| ≥65 | -1.86 (2.29) | 0.417 | -0.025 (0.028) | 0.367 | 1.7 (5.5) | 0.760 | 0.3 (7.4) | 0.964 | -1.4 (7.9) | 0.863 | 6.9 (4.7) | 0.139 |
| Gender |  |  |  |  |  |  |  |  |  |  |  |  |
| Male | 4.07 (1.30) | 0.002 | 0.028 (0.016) | 0.080 | -2.1 (2.2) | 0.360 | -6.8 (2.9) | 0.019 | -5.8 (3.1) | 0.057 | -9.8 (2.6) | <0.001 |
| Female | Reference |  | Reference |  | Reference |  | Reference |  | Reference |  | Reference |  |
| Race / Ethnicity |  |  |  |  |  |  |  |  |  |  |  |  |
| White (non-Hispanic) | Reference |  | Reference |  | Reference |  | Reference |  | Reference |  | Reference |  |
| Black/African<br>American | 2.17 (2.73) | 0.428 | 0.017 (0.033) | 0.613 | -1.8 (5.1) | 0.726 | 0.8 (6.4) | 0.899 | 0.1 (7.1) | 0.994 | -1.9 (5.6) | 0.730 |
| Hispanic or Latino | 0.65 (1.68) | 0.701 | 0.005 (0.021) | 0.818 | -1.9 (3.0) | 0.530 | 4.3 (3.9) | 0.268 | 4.8 (4.1) | 0.243 | -2.3 (3.4) | 0.494 |
| Asian | 4.24 (2.54) | 0.096 | 0.032 (0.031) | 0.316 | -1.7 (4.7) | 0.713 | 1.4 (5.8) | 0.807 | 2.4 (6.3) | 0.705 | 0.9 (5.2) | 0.858 |
| Patient refused | -0.30 (3.26) | 0.927 | 0.031 (0.040) | 0.438 | -3.3 (6.1) | 0.589 | 1.1 (8.4) | 0.897 | -2.3 (9.0) | 0.800 | -6.4 (7.0) | 0.359 |
| Other | 2.90 (2.81) | 0.303 | 0.019 (0.035) | 0.582 | 3.9 (4.8) | 0.418 | -6.9 (6.5) | 0.288 | -3.1 (6.9) | 0.661 | -7.5 (5.8) | 0.198 |
| Region |  |  |  |  |  |  |  |  |  |  |  |  |
| Northeast | 1.91 (2.00) | 0.341 | 0.015 (0.025) | 0.534 | 6.2 (3.3) | 0.057 | -1.9 (4.2) | 0.648 | -0.5 (4.5) | 0.908 | -2.3 (3.9) | 0.551 |
| South | 0.87 (1.51) | 0.568 | -0.002 (0.019) | 0.905 | -1.0 (2.6) | 0.698 | -2.4 (3.3) | 0.466 | -2.4 (3.5) | 0.507 | -2.9 (3.0) | 0.328 |
| Midwest | Reference |  | Reference |  | Reference |  | Reference |  | Reference |  | Reference |  |
| West | 0.49 (2.08) | 0.815 | 0.019 (0.026) | 0.448 | -0.7 (3.4) | 0.830 | -0.8 (4.5) | 0.857 | -1.3 (4.8) | 0.793 | -3.7 (4.2) | 0.374 |
| Social Vulnerability Index |  |  |  |  |  |  |  |  |  |  |  |  |
| <0.25 | Reference |  | Reference |  | Reference |  | Reference |  | Reference |  | Reference |  |
| ≥0.25 and <0.5 | 1.21 (1.45) | 0.403 | 0.034 (0.018) | 0.060 | 1.3 (2.4) | 0.579 | -3.9 (3.0) | 0.196 | -4.6 (3.3) | 0.158 | -5.2 (2.9) | 0.076 |
| ≥0.5 and <0.75 | 0.93 (1.62) | 0.568 | 0.020 (0.020) | 0.321 | 4.7 (2.8) | 0.090 | -4.8 (3.6) | 0.180 | -6.0 (3.9) | 0.124 | -3.6 (3.3) | 0.276 |
| ≥0.75 | -2.80 (2.14) | 0.192 | 0.009 (0.026) | 0.740 | 2.3 (3.7) | 0.531 | -9.3 (4.7) | 0.048 | -7.8 (5.0) | 0.121 | -5.0 (4.3) | 0.247 |
| Previously tested positive | 0.99 (1.13) | 0.381 | 0.003 (0.014) | 0.816 | -1.9 (2.0) | 0.333 | -0.5 (2.6) | 0.851 | -2.7 (2.8) | 0.329 | -1.2 (2.3) | 0.591 |
| Work in healthcare | -0.90 (1.80) | 0.616 | -0.018 (0.022) | 0.424 | 3.1 (2.8) | 0.272 | 0.8 (3.6) | 0.818 | -0.7 (4.0) | 0.851 | 3.7 (3.6) | 0.300 |
| Work in high-risk setting | -2.83 (1.96) | 0.150 | -0.006 (0.024) | 0.790 | 4.7 (3.4) | 0.166 | 10.3 (4.3) | 0.017 | 13.3 (4.6) | 0.004 | 5.7 (4.0) | 0.155 |
| Live in high-risk setting | 1.36 (2.53) | 0.591 | -0.028 (0.031) | 0.370 | 3.2 (4.7) | 0.502 | 4.8 (6.0) | 0.431 | 7.2 (6.5) | 0.268 | 12.1 (5.0) | 0.017 |
| Immunocompromised | 1.22 (2.61) | 0.639 | -0.018 (0.032) | 0.560 | 13.1 (6.1) | 0.033 | 9.9 (8.4) | 0.243 | 7.9 (9.1) | 0.385 | 5.5 (5.1) | 0.282 |
| Number of Symptoms on index day | -0.59 (0.23) | 0.009 | -0.010 (0.003) | 0.001 | 0.8 (0.4) | 0.048 | 1.3 (0.5) | 0.008 | 1.9 (0.5) | 0.001 | 1.8 (0.4) | <0.001 |

**Supplemental Figure 1.**
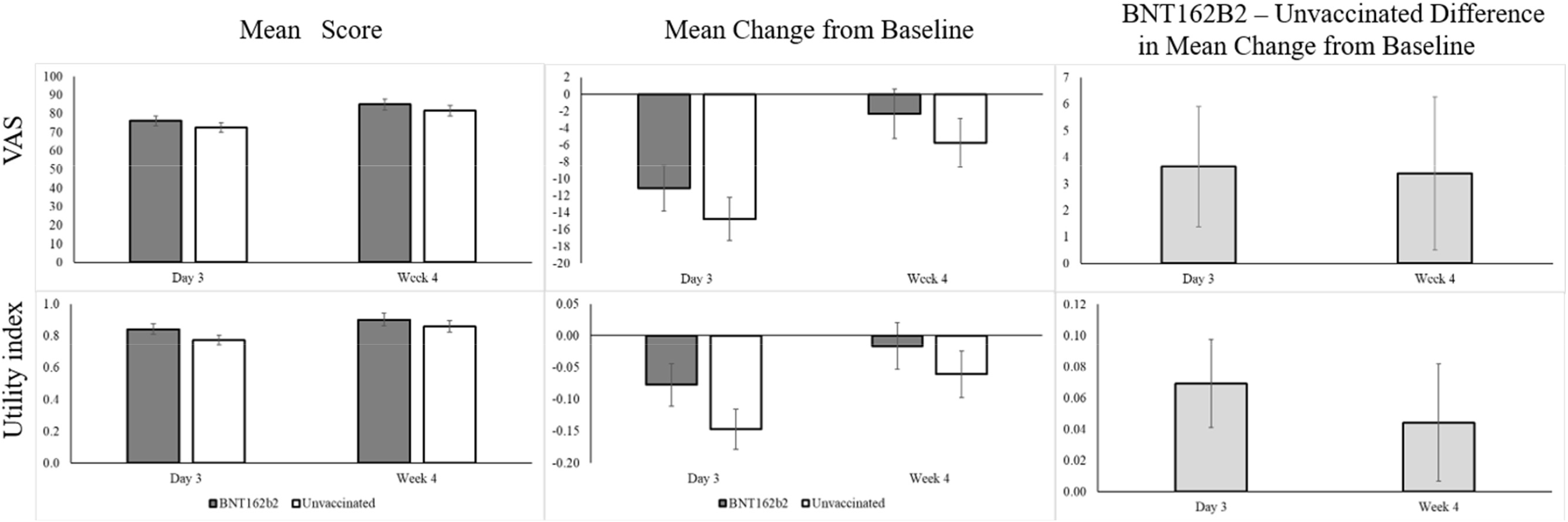
Least-Square Estimates and 95% Confidence Intervals of EQ-5D-5L Scores ^a^ ^**a**^ Score ranges: EQ-5D-5L VAS 0 to 100, EQ-5D-5L UI (the United States weights) −0.573 to 1.

**Supplemental Figure 2.**
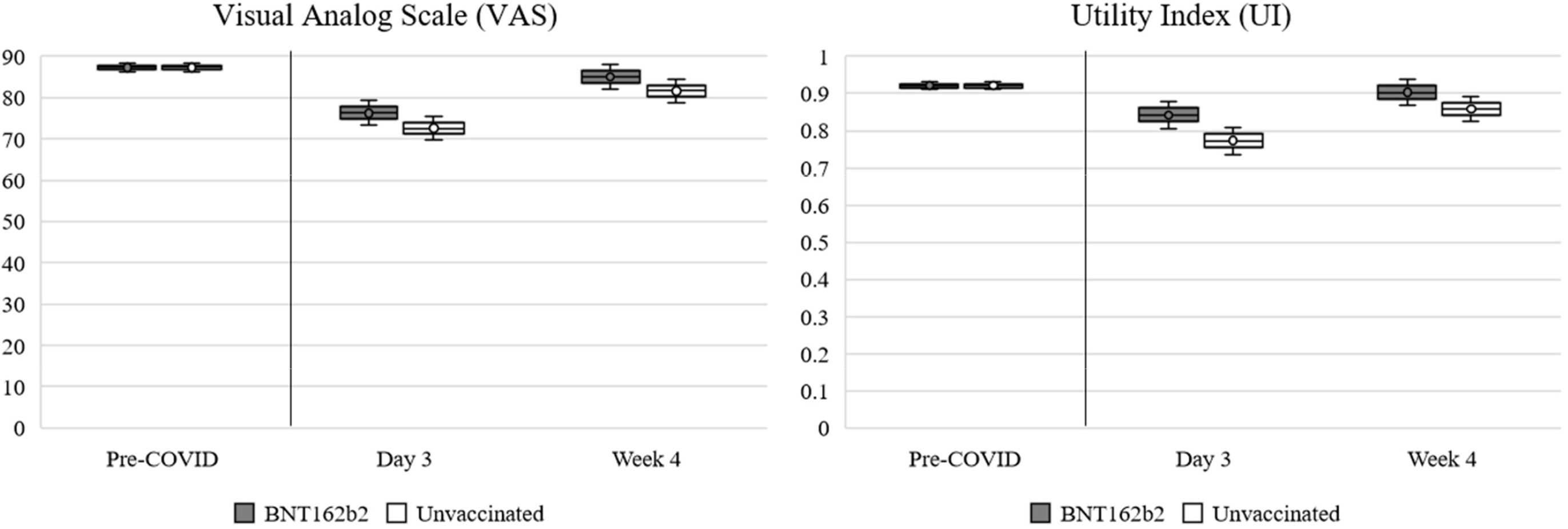
Summary Results of EQ-5D-5L scores across time periods ^a^ ^**a**^ Score ranges: EQ-5D-5L VAS 0 to 100, EQ-5D-5L UI (the United States weights) −0.573 to 1. Dots are the mean values and whiskers are the 95% Confidence Intervals. The Pre-COVID values represent the pooled means with 95% Confidence Internals. The Day 3 and Week 4 values are the least square estimate scores.

**Supplemental Figure 3.**
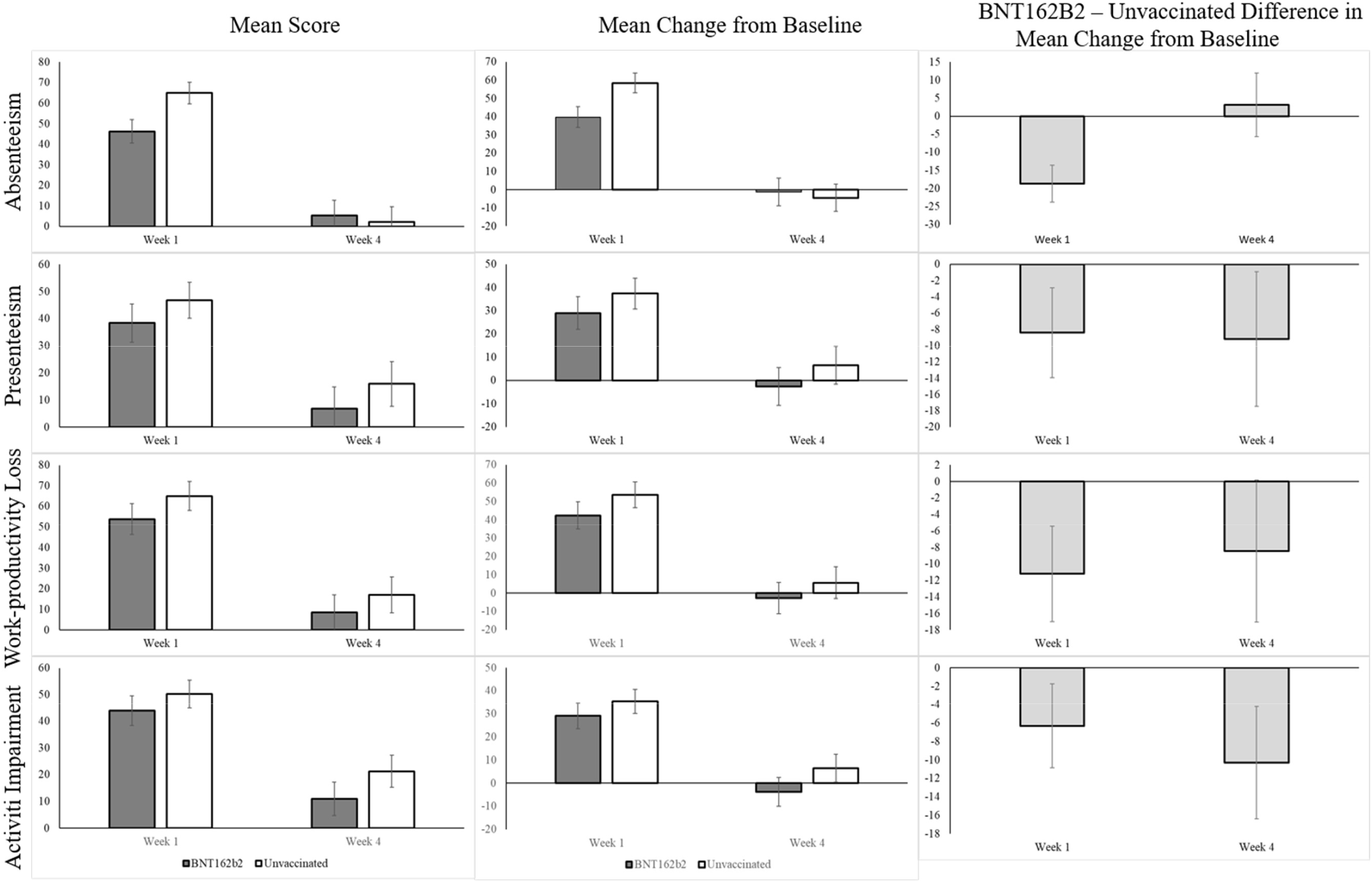
Least-Square Estimates and 95% Confidence Intervals of WPAI-GH Scores ^a^ ^a^ WPAI-GH score (absenteeism, presenteeism, work productivity loss, and activity impairment) ranges from 0 to 100 percent.

**Supplemental Figure 4.**
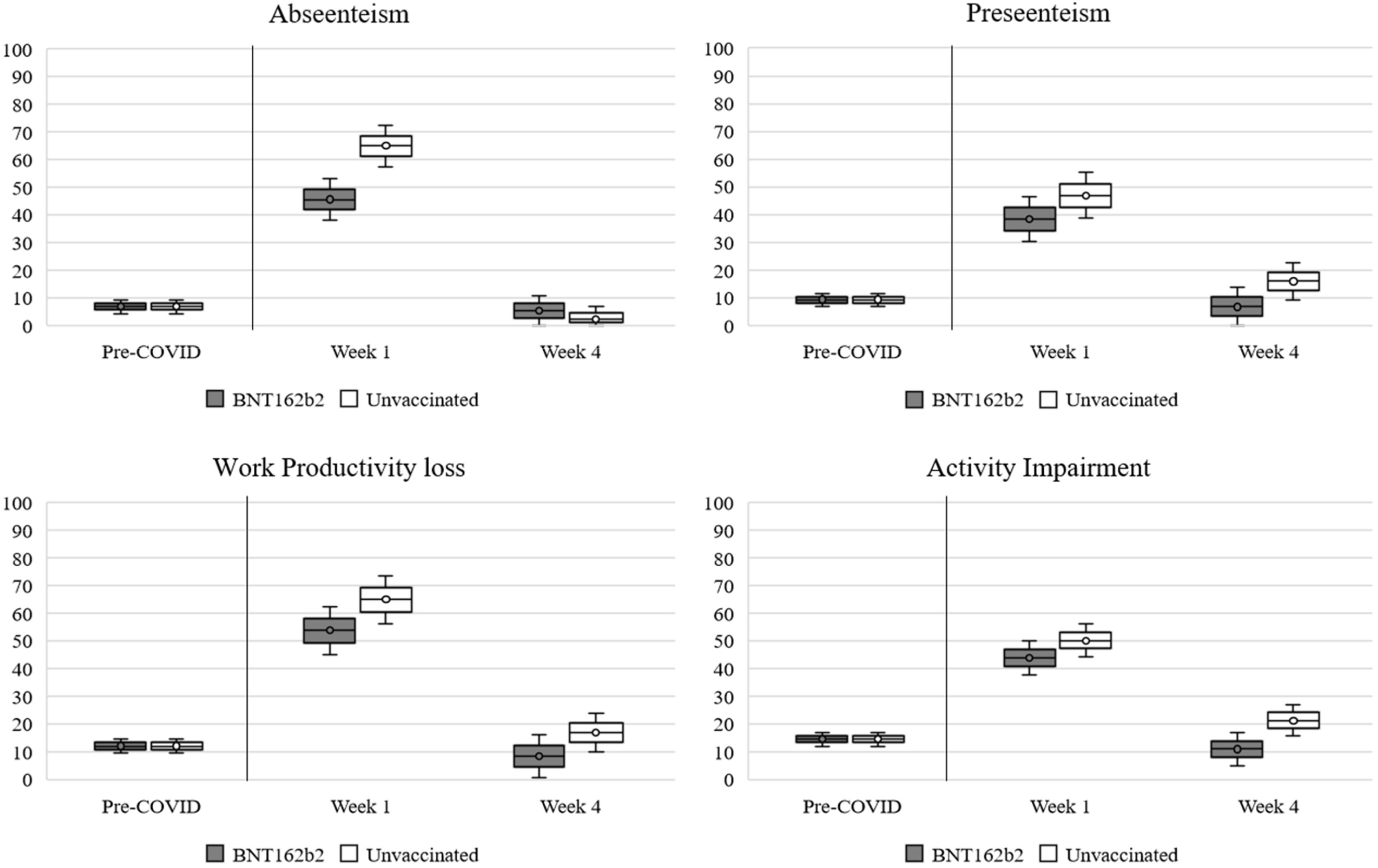
Summary Results of WPAI-GH scores across time periods ^a^ ^a^ WPAI-GH score (absenteeism, presenteeism, work productivity loss, and activity impairment) ranges from 0 to 100 percent. Dots are the mean values and whiskers are the 95% Confidence Intervals. The Pre-COVID values represent the pooled means with 95% Confidence Internals. The Week 1 and Week 4 values are the least square estimate scores.

**Supplemental Table 4.** Model Predicting the Missingness at Week 1 and Week 4

|  | Summary: % (n) / Mean (SD) |  |  | Model |  |  |
| --- | --- | --- | --- | --- | --- | --- |
|  | Total | Week 1 Missing | Week 4 Missing | Coeff (SE) | P value | Odds Ratio |
| Intercept | 430 | 11.9% (51) | 23.0% (99) | -3.14 (1.19) | 0.009 |  |
| Assessment Time |  |  |  |  |  |  |
| Week 4 | 430 |  | 23.0% (99) | 0.85 (0.12) | <0.001 | 2.3 (1.9, 2.9) |
| Week 1 | 430 | 11.9% (51) |  | Reference | . | 1.0 |
| Vaccination status |  |  |  |  |  |  |
| BNT162b2 | 233 | 12.9% (30) | 24.5% (57) | 0.36 (0.26) | 0.166 | 1.4 (0.9, 2.4) |
| Unvaccinated | 197 | 10.7% (21) | 21.3% (42) | Reference | . | 1.0 |
| EQ VAS on Day 3 | 72.9<br>(17.5) | 68.9 (19.1) | 71.7 (17.3) | 0.00 (0.01) | 0.967 | 1.0 (1.0, 1.0) |
| EQ VAS change from pre-COVID-19 baseline to Day 3 | -14.4<br>(14.8) | -18.2 (18.9) | -16.0 (16.6) | -0.01 (0.01) | 0.311 | 1.0 (1.0, 1.0) |
| Age, years |  |  |  |  |  |  |
| 18-29 | 87 | 8.0% (7) | 16.1% (14) | Reference | . | 1.0 |
| 30-49 | 213 | 10.3% (22) | 23.9% (51) | 0.69 (0.36) | 0.052 | 2.0 (1.0, 4.0) |
| 50-64 | 94 | 16.0% (15) | 27.7% (26) | 0.94 (0.40) | 0.019 | 2.6 (1.2, 5.6) |
| ≥65 | 36 | 19.4% (7) | 22.2% (8) | 0.72 (0.52) | 0.161 | 2.1 (0.8, 5.6) |
| Gender |  |  |  |  |  |  |
| Male | 103 | 4.9% (5) | 16.5% (17) | -0.80 (0.32) | 0.013 | 0.4 (0.2, 0.8) |
| Female | 327 | 14.1% (46) | 25.1% (82) | Reference | . | 1.0 |
| Race / Ethnicity |  |  |  |  |  |  |
| White (non-Hispanic) | 295 | 9.8% (29) | 19.7% (58) | Reference | . | 1.0 |
| Black/African American | 20 | 20.0% (4) | 35.0% (7) | 0.67 (0.50) | 0.184 | 2.0 (0.7, 5.3) |
| Hispanic or Latino | 61 | 11.5% (7) | 27.9% (17) | 0.21 (0.36) | 0.565 | 1.2 (0.6, 2.5) |
| Asian | 22 | 18.2% (4) | 27.3% (6) | 0.37 (0.52) | 0.480 | 1.4 (0.5, 4.0) |
| Patient refused | 13 | 30.8% (4) | 30.8% (4) | 0.94 (0.62) | 0.128 | 2.6 (0.8, 8.6) |
| Other | 19 | 15.8% (3) | 36.8% (7) | 1.22 (0.54) | 0.024 | 3.4 (1.2, 9.8) |
| Region |  |  |  |  |  |  |
| Northeast | 52 | 7.7% (4) | 21.2% (11) | 0.50 (0.48) | 0.295 | 1.7 (0.6, 4.2) |
| South | 252 | 13.5% (34) | 24.6% (62) | 0.32 (0.37) | 0.378 | 1.4 (0.7, 2.8) |
| Midwest | 77 | 6.5% (5) | 16.9% (13) | Reference | . | 1.0 |
| West | 49 | 16.3% (8) | 26.5% (13) | 0.44 (0.47) | 0.343 | 1.6 (0.6, 3.9) |
| Social Vulnerability Index |  |  |  |  |  |  |
| <0.25 | 98 | 12.2% (12) | 22.4% (22) | Reference | . | 1.0 |
| ≥0.25 and <0.5 | 164 | 9.8% (16) | 19.5% (32) | -0.22 (0.33) | 0.496 | 0.8 (0.4, 1.5) |
| ≥0.5 and <0.75 | 120 | 12.5% (15) | 25.0% (30) | 0.07 (0.35) | 0.850 | 1.1 (0.5, 2.1) |
| ≥0.75 | 48 | 16.7% (8) | 31.3% (15) | 0.38 (0.45) | 0.397 | 1.5 (0.6, 3.5) |
| Previously tested positive | 167 | 15.6% (26) | 26.9% (45) | 0.45 (0.25) | 0.069 | 1.6 (1.0, 2.5) |
| Work in healthcare | 47 | 12.8% (6) | 19.1% (9) | -0.24 (0.42) | 0.567 | 0.8 (0.3, 1.8) |
| Work in high-risk setting | 44 | 13.6% (6) | 22.7% (10) | -0.25 (0.43) | 0.559 | 0.8 (0.3, 1.8) |
| Live in high-risk setting | 22 | 9.1% (2) | 27.3% (6) | 0.38 (0.54) | 0.484 | 1.5 (0.5, 4.3) |
| Immunocompromised | 19 | 10.5% (2) | 15.8% (3) | -0.33 (0.67) | 0.623 | 0.7 (0.2, 2.7) |
| Number of Symptoms on index day | 5.3 (2.6) | 5.4 (2.6) | 5.0 (2.5) | -0.07 (0.05) | 0.188 | 0.9 (0.8, 1.0) |

## REFERENCES

1. Poudel AN, Zhu S, Cooper N, Roderick P, Alwan N, Tarrant C, Ziauddeen N, Yao GL (2021) Impact of Covid-19 on health-related quality of life of patients: A structured review. PLoS One. 16(10):e0259164.

2. Nandasena H, Pathirathna M, Atapattu A, Prasanga P (2022) Quality of life of COVID 19 patients after discharge: Systematic review. PloS one. 17(2):e0263941.

3. Figueiredo EAB, Silva WT, Tsopanoglou SP, Vitorino DFdM, Oliveira LFLd, Silva KLS, Luz HDH, Ávila MR, Oliveira LFFd, Lacerda ACR (2022) The health-related quality of life in patients with post-COVID-19 after hospitalization: a systematic review. Revista da Sociedade Brasileira de Medicina Tropical. 55.

4. Amdal CD, Pe M, Falk RS, Piccinin C, Bottomley A, Arraras JI, Darlington AS, Hofsø K, Holzner B, Jørgensen Nmh (2021) Health-related quality of life issues, including symptoms, in patients with active COVID-19 or post COVID-19; a systematic literature review. Quality of Life Research. 30(12):3367–81.

5. International Vaccine Access Center, Johns Hopkins Bloomberg School of Public Health, World Health Organization. Results of COVID-19 Vaccine Effectiveness & Impact Studies: An Ongoing Systematic Review. https://view-hub.org/sites/default/files/2022-04/COVID19_VE_and_Impact_Lit_Review_Methods.pdf Accessed August 15, 2022.

6. Arab-Zozani M, Hashemi F, Safari H, Yousefi M, Ameri H (2020) Health-related quality of life and its associated factors in COVID-19 patients. Osong public health and research perspectives. 11(5):296.

7. Petersen EL, Goßling A, Adam G, Aepfelbacher M, Behrendt C-A, Cavus E, Cheng B, Fischer N, Gallinat J, Kühn S (2022) Multi-organ assessment in mainly non-hospitalized individuals after SARS-CoV-2 infection: the Hamburg City Health Study COVID programme. European Heart Journal. 43(11):1124–37.

8. McKay SC, Lembach H, Hann A, Okoth K, Anderton J, Nirantharakumar K, Magill L, Torlinska B, Armstrong M, Mascaro J (2021) Health-related quality of life, uncertainty and coping strategies in solid organ transplant recipients during shielding for the COVID-19 pandemic. Transplant International. 34(11):2122–37.

9. Talman S, Boonman-de Winter L, De Mol M, Hoefman E, Van Etten R, De Backer I (2021) Pulmonary function and health-related quality of life after COVID-19 pneumonia. Respiratory Medicine. 176:106272.

10. Centers for Disease Control and Prevention. Updates - Symptoms of COVID-19. https://www.cdc.gov/coronavirus/2019-ncov/symptoms-testing/symptoms.html Accessed August 1 2021.

11. EuroQol Research Foundation. (2019) EQ-5D-5L User Guide, Version 3.0. https://euroqol.org/publications/user-guides Accessed August 5, 2021.

12. Cha AS, Law EH, Shaw JW, Pickard AS (2019) A comparison of self-rated health using EQ-5D VAS in the United States in 2002 and 2017. Quality of Life Research. 28(11):3065–9.

13. Pickard AS, Law EH, Jiang R, Pullenayegum E, Shaw JW, Xie F, Oppe M, Boye KS, Chapman RH, Gong CL (2019) United States valuation of EQ-5D-5L health states using an international protocol. Value in Health. 22(8):931–41.

14. Reilly MC, Zbrozek AS, Dukes EM (1993) The validity and reproducibility of a work productivity and activity impairment instrument. Pharmacoeconomics. 4(5):353–65.

15. Reilly Associates. (2002) WPAI Scoring. http://www.reillyassociates.net/WPAI_Scoring.html Accessed August 5, 2021.

16. Centers for Disease Control and Prevention. Long COVID or Post-COVID Conditions. https://www.cdc.gov/coronavirus/2019-ncov/long-term-effects/index.html Accessed August 15, 2022.

17. Rosner B (2015) Fundamentals of biostatistics. Eighth ed. Cengage learning, Boston, MA

18. Freeman G, Halton JH (1951) Note on an exact treatment of contingency, goodness of fit and other problems of significance. Biometrika. 38(1/2):141–9.

19. Fitzmaurice GM, Laird NM, Ware JH (2012) Applied longitudinal analysis. John Wiley & Sons, Hoboken, NJ

20. Cohen J (1988) Statistical power analysis for the behavioral sciences. 2nd ed. Lawrence Erlbaum Assoc, Hillsdale, NJ.

21. McLeod LD, Cappelleri JC, Hays RD (2016) Best (but oft-forgotten) practices: expressing and interpreting associations and effect sizes in clinical outcome assessments. The American journal of clinical nutrition. 103(3):685–93. Erratum: https://doi.org/10.3945/ajcn.116.148593.

22. Mouelhi Y, Jouve E, Castelli C, Gentile S (2020) How is the minimal clinically important difference established in health-related quality of life instruments? Review of anchors and methods. Health and Quality of Life Outcomes. 18(1):1–17.

23. STROBE Statement—Checklist of items that should be included in reports of cohort studies. https://www.strobe-statement.org/download/strobe-checklist-cohort-studies-pdf Accessed August 5, 2022.

24. Jacobson KB, Rao M, Bonilla H, Subramanian A, Hack I, Madrigal M, Singh U, Jagannathan P, Grant P (2021) Patients with uncomplicated COVID-19 have long-term persistent symptoms and functional impairment similar to patients with severe COVID-19: a cautionary tale during a global pandemic. Clinical Infectious Diseases. 10.1093/cid/ciab103.

25. Hernandez-Romieu AC, Leung S, Mbanya A, Jackson BR, Cope JR, Bushman D, Dixon M, Brown J, McLeod T, Saydah S (2021) Health care utilization and clinical characteristics of nonhospitalized adults in an integrated health care system 28–180 days after COVID-19 diagnosis—Georgia, May 2020–March 2021. Morbidity and Mortality Weekly Report. 70(17):644.

26. Jiang R, Janssen M, Pickard AS (2021) US population norms for the EQ-5D-5L and comparison of norms from face-to-face and online samples. Quality of Life Research. 30(3):803–16.

27. Lawson A, Tan AC, Naylor J, Harris IA (2020) Is retrospective assessment of health-related quality of life valid? BMC Musculoskeletal Disorders. 21(1):1–10.

28. Rajan SS, Wang M, Singh N, Jacob AP, Parker SA, Czap AL, Bowry R, Grotta JC, Yamal JM (2021) Retrospectively Collected EQ-5D-5L Data as Valid Proxies for Imputing Missing Information in Longitudinal Studies. Value Health. 24(12):1720–7. 10.1016/j.jval.2021.07.007.

29. Tundia N, Hass S, Fuldeore M, Wang LL, Cavanaugh T, Boone J, Heaton P (2015) Validation and US population norms of health-related productivity questionnaire. Value in Health. 18(3):A24.

30. Romano C, Fehnel S, Stoddard J, Sadoff J, Lewis S, McNulty P, Chan EK, Evans E, Jamieson C, Slagle AF (2022) Development of a novel patient-reported outcome measure to assess signs and symptoms of COVID-19. Journal of Patient-Reported Outcomes. 6(1):1–12.

31. Hughes SE, Haroon S, Subramanian A, McMullan C, Aiyegbusi OL, Turner GM, Jackson L, Davies EH, Frost C, McNamara G (2022) Development and validation of the symptom burden questionnaire for long covid (SBQ-LC): Rasch analysis. BMJ. 377.

